# Metabolomics reveals reasons for the efficacy of acupuncture in migraine patients: The role of anaerobic glycolysis and mitochondrial citrate in migraine relief

**DOI:** 10.1101/2023.10.12.22271400

**Authors:** Zishan Gao, Xianzhong Yan, Rui Wang-Sattler, Marcela Covic, Guang Yu, Feifei Ge, Jia Lin, Qin Chen, Juan Liu, Sapna Sharma, Sophie Molnos, Brigitte Kuehnel, Rory Wilson, Jonathan Adam, Stefan Brandmaier, Shuguang Yu, Ulrich Mansmann, Fanrong Liang, Christian Gieger

## Abstract

Acupuncture is used worldwide to treat migraine, but its scientific mechanism remains unclear. Here, we report a 1H NMR metabolomics study involving 40 migraine patients and 10 healthy people randomly receiving acupuncture or sham acupuncture, followed by machine learning techniques and functional analysis. We found that acupuncture at acupoints particularly enhanced anaerobic glycolysis and modified mitochondrial function by adjusting the levels of plasma pyruvic acid (P = 0.012), lactic acid (P = 0.031) and citrate (P = 0.00079) at a Bonferroni-corrected level of significance compared to the pre-treatment level of these three metabolites in migraine patients. Therefore, acupuncture supplies energy to migraine patients and relieves migraine attacks. In contrast, we observed that sham acupuncture may partially supply energy to migraine patients through lipid metabolism by changing the levels of plasma lipid (P = 0.0012), glycerine (P = 0.021), and pyruvic acid (P = 0.047) at a Bonferroni-corrected level of significance. The functional network analysis further indicates this different way of supplying energy contributes to the different effects of acupuncture and sham acupuncture. Our findings reveal novel metabolic evidence for the specific effect of acupuncture in relation to sham acupuncture. This metabolic evidence could enlighten a brand new direction into acupuncture analgesia mechanism, which in turn would pose fresh challenges for future acupuncture research.

## Introduction

Acupuncture is commonly used for preventing and relieving migraine worldwide (Wells *et al*.,2011). Understanding the mechanism underlying the efficacy of acupuncture for migraine is key to the acceptance of acupuncture as valid therapy for both Western doctors and policymakers. Recently, a rich source of clinical evidence has demonstrated the effectiveness and safety of acupuncture for migraine, showing that acupuncture could effectively reduce the intensity of migraine, frequency of migraine attacks, and number of migraine days (Linde *et al*., 2016; Vickers *et al*., 2012). In contrast to the abundant evidence from those clinical trials, the biological basis for the efficacy of acupuncture in relieving migraines remains unclear.

To date, several experimental studies have shown that acupuncture alleviates migraine by activating a range of biochemicals in peripheral and pain-related central nuclei (Goldman *et al*., 2010; Zhao *et al*.,2017). Goldman et al (2010) found that adenosine and adenosine metabolism mediated the analgesic effect of acupuncture in a mouse model. Zhao et al (2017) showed that calcitonin gene-related peptide (CGRP), which plays a key role in triggering migraine, was suppressed by electroacupuncture in a rat model of migraine. However, there is no validated biomarker associated with the effects of acupuncture for migraine in clinical studies. The explicit biochemical pathways or mechanisms addressing the process of acupuncture relief of migraine are not clear at present. Further, the specific effect of acupuncture relative to sham acupuncture is a long-standing controversial topic. Zhao L et al showed in a multicentre randomized trial that acupuncture for migraine is more effective than sham acupuncture (Zhao *et al*., 2017); others, however, have found no differences (Linde *et al*., 2005; Li *et al*., 2012). An individual patient data (IPD) meta-analysis including 20,827 patients confirmed that acupuncture is statistically superior to sham acupuncture for migraine, but the difference in effect size between acupuncture and sham was relatively small (Vickers *et al*., 2012; Vickers *et al*., 2014). Due to the absence of explainable biological mechanisms for the specific effects of acupuncture, the acceptance of acupuncture as a referable therapy in migraine management is still arguable (Linde *et al*., 2017). Hence, two important questions are raised in this paper: first, are there any biomarkers or pathways that provide a possible mechanism for the efficacy of acupuncture relieving migraine? Second, is there any biological basis attributed to the specific effect of acupuncture compared with sham acupuncture for relieving migraine?

Recently, a large number of studies resumed investigation of the pathogenesis of migraine, emphasizing that abnormalities in energy metabolism and mitochondrial function are the fundamental milestones in the pathophysiology of migraine (Colombo *et al*., 2014; Lodi *et al*.,2006; Sparaco *et al*., 2006). Increasing experiments have revealed that insufficient cerebral glycogen unbalances energy metabolism, leading to inhibition of astroglial mitochondrial respiration and excessive production of free radicals. These metabolic changes are attributed to further energy failure in neurons that stimulate cortical expansion depression (CSD) and thus trigger migraine (Finsterer *et al*., 2018). Moreover, previous studies indicated that the effect of acupuncture may involve multiple pathways and dynamic system changes from genomics to metabolomics (Xu *et al*., 2012). Metabolomics has been powerfully utilized to reveal potential metabolic biomarkers of acupuncture for hypertension (Yang *et al*., 2018), functional dyspepsia (Wu *et al*., 2016) and chronic atrophic gastritis (Luo *et al*., 2013). As such, it is therefore essential to employ metabolomic approaches to pinpoint crucial biomarkers and elucidate the systemic mechanism of acupuncture effects for migraine. Our previous study demonstrated that acupuncture could adjust metabolic profiling in an acute migraine rat model (Gao *et al*., 2014). Nevertheless, the challenge from statistical methods for dealing with high-dimensional metabolomic data still limits the use of metabolomics in acupuncture research. Within this context, machine learning techniques, such as the least absolute shrinkage and selection operator (Lasso), are promising tools for reducing high-dimensional data and selecting accurate biomarkers for metabolomics (LeWitt *et al*., 2017; Menni *et al*., 2017).

To investigate the potential metabolic mechanism for the efficacy of acupuncture relief of migraine, we employed ^1^H-Nuclear magnetic resonance (NMR) metabolomic technology to detect plasma metabolic phenotypes for 40 female migraine patients who randomly received either acupuncture or sham acupuncture from a total of 476 patients in a randomized controlled trial, together with a group of 10 healthy persons. Using orthogonal signal correction for partial least square discriminate analysis (OPLS-DA), we determined different metabolic profiles among healthy controls and migraine patients treated with acupuncture or sham acupuncture. Next, we conducted Lasso regression to select metabolic biomarkers for discriminating acupuncture and sham acupuncture and validated the selected metabolites using analysis of variance (ANOVA) combined with Box-Cox transformations and Bonferroni corrections. Further, we validated the performance of Bonferroni-corrected significant metabolic biomarkers discriminating migraine patients at baseline, healthy people and migraine patients after acupuncture in a clinical setting by receiver operating characteristic (ROC) curve analysis. Finally, we complemented our analysis with 2 Bonferroni-corrected significant metabolites and 2 discriminated metabolic pathways attributed to the specific effect of acupuncture using Ingenuity Pathway Analysis (IPA), revealing a novel metabolic mechanism for the efficacy of acupuncture in relieving migraine (Figure 1).

**Fig. 1.**
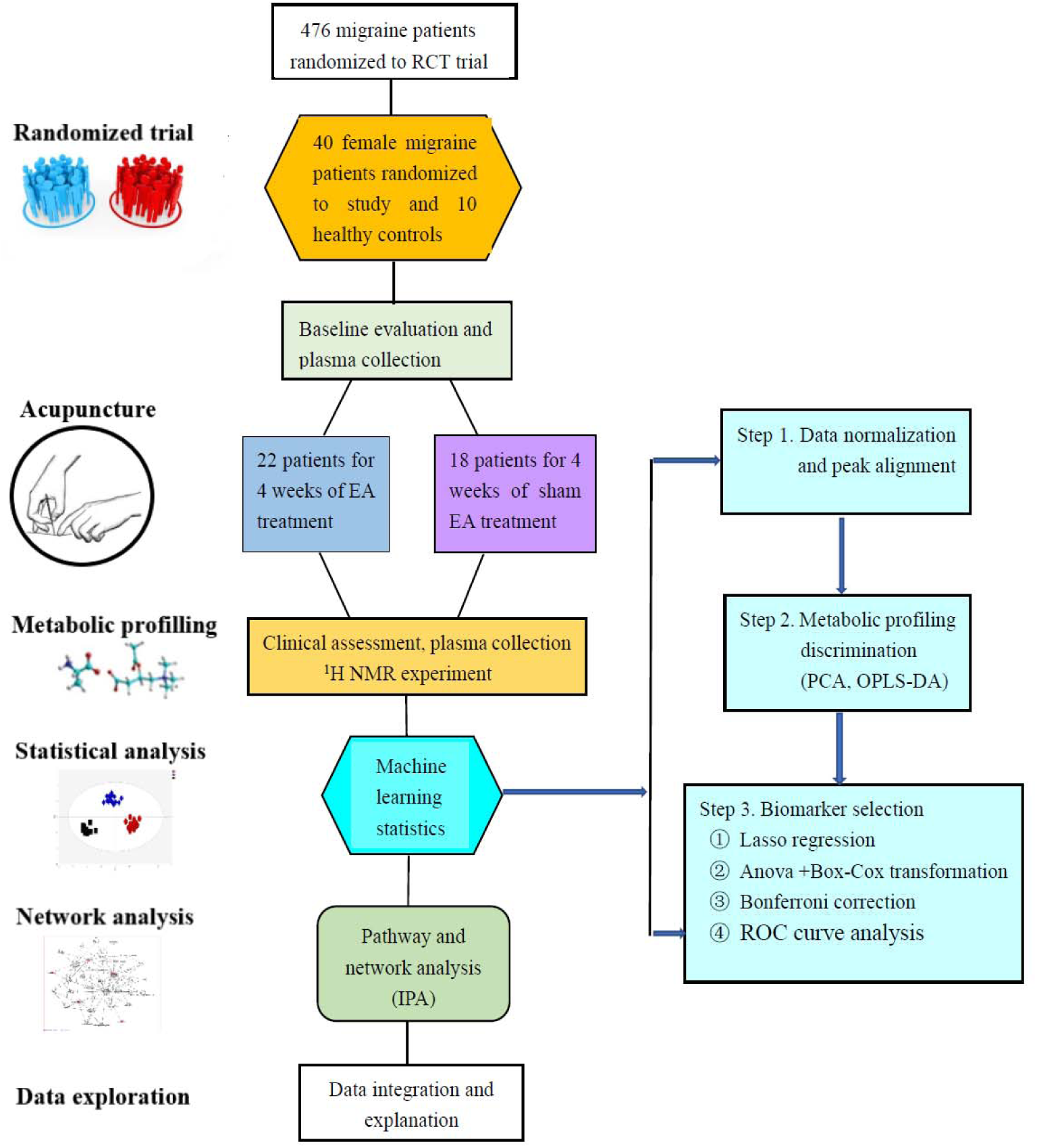
Study design. We conducted a non-targeted metabolomic study of 40 migraine patients and 10 healthy people. Plasma was collected before and after EA or Sham EA treatment for ^1^H NMR experiments.

## Results

### Baseline characteristics and clinical effects of acupuncture

A total of 476 migraine patients were included in the multicenter trial (Li *et al*., 2012), and 40 eligible female patients were enrolled in the 4-week baseline period and randomized into the true electroacupuncture (EA) and sham electroacupuncture groups (Sham EA) (22 in the EA group, 18 in the Sham EA group) of the metabolomic study. These 40 patients finished the 4-week baseline assessment and the 4-week EA or Sham EA treatment (Figure 1). There are 39 patients with migraine who meet the definition of migraine without aura, only one migraine patient with aura was recruited in the study. And this patient dropped out in the end of electroacupuncture treatment and not included in the statistical analysis. The baseline characteristics and clinical outcomes were based on the intention-to-treat (ITT) population in this study. We omitted the cases that retained only the baseline measurement but had missing data in all clinical outcomes. No significant difference between the two groups was found for any demographic characteristics of all included patients, including age, sex, height, weight, or disease status of the patients, such as duration of disease, number of days with migraine, and visual analogue scale (VAS) that assesses pain severity (Table 1, Table S1).

**Table 1.**
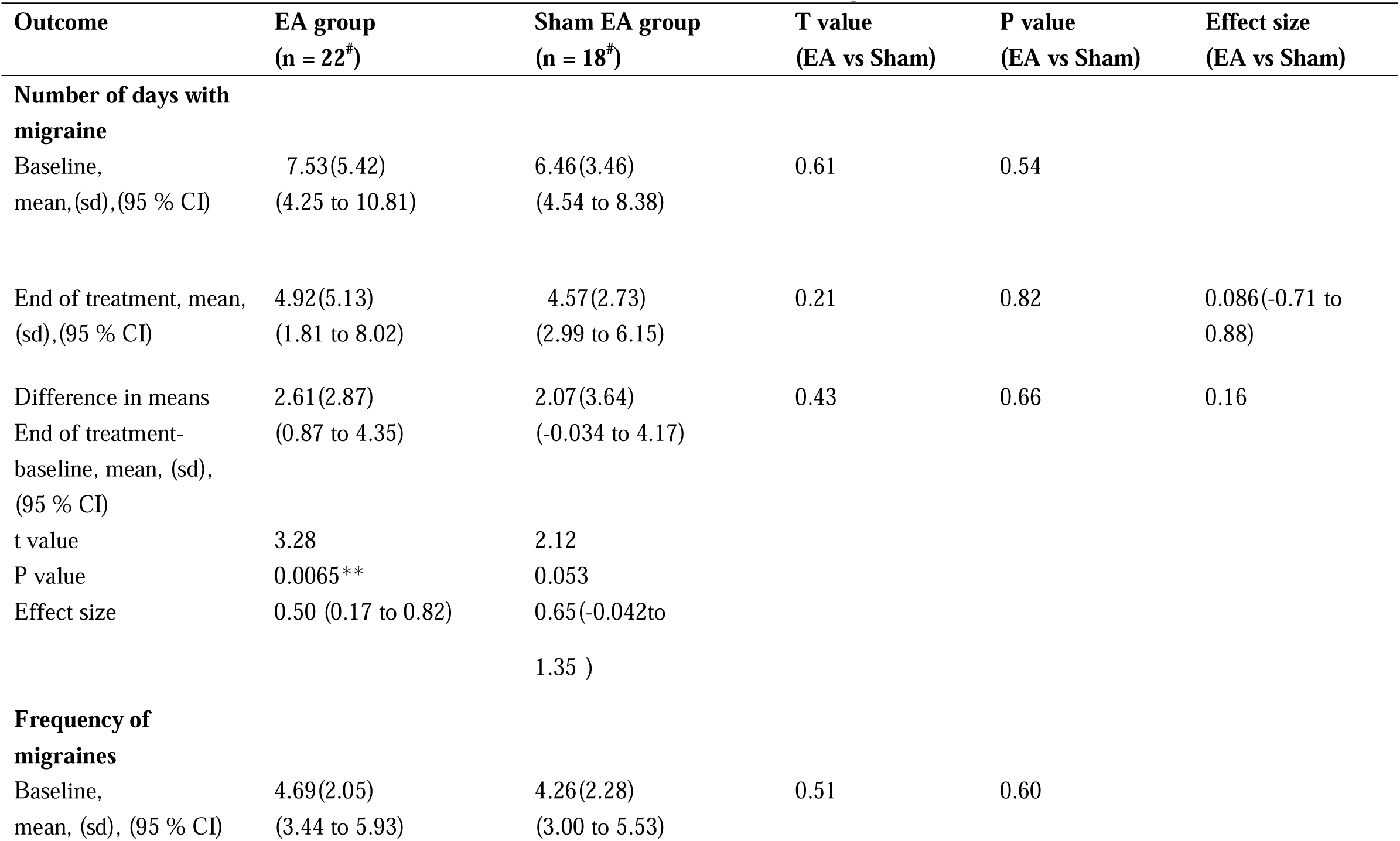
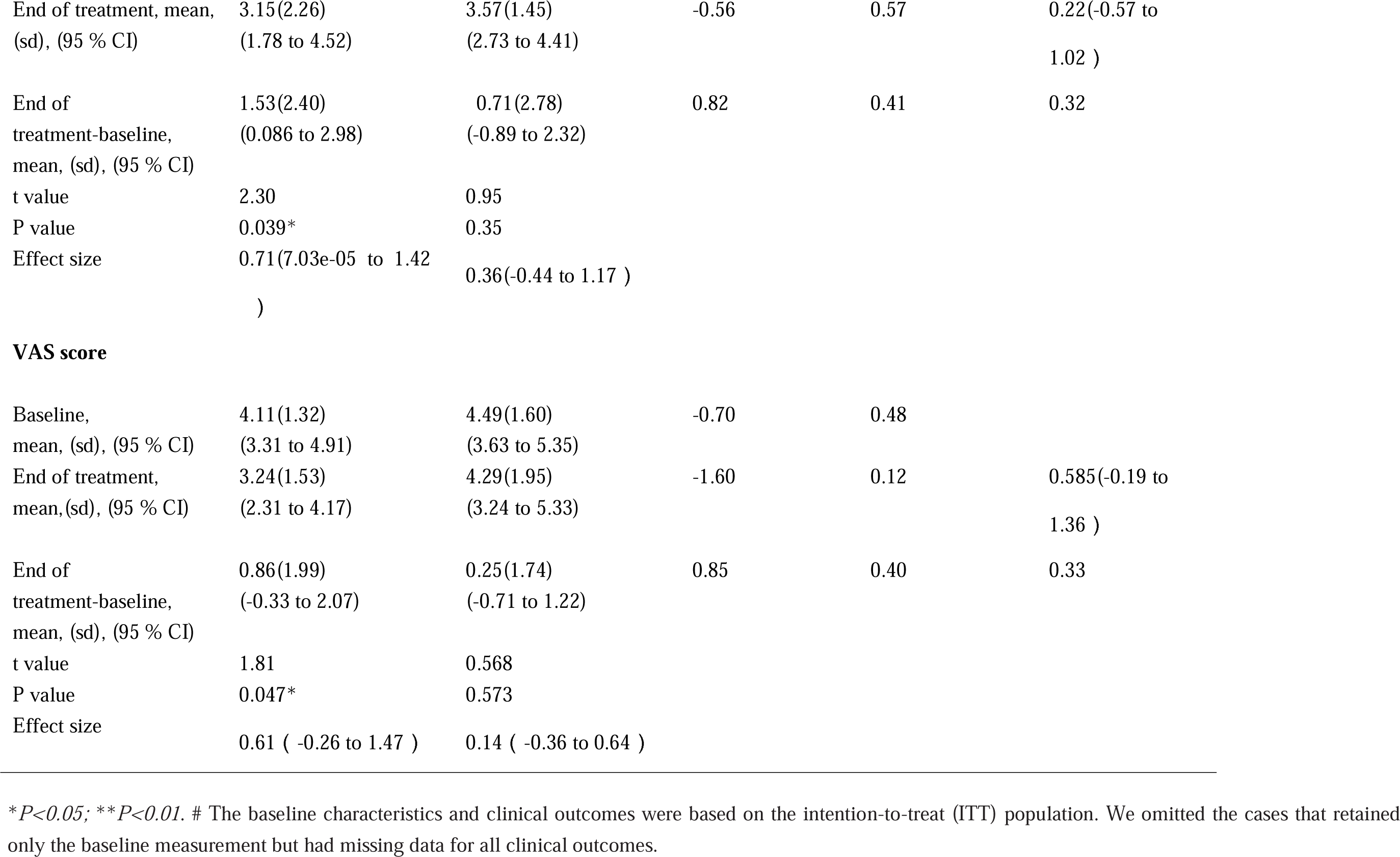
Clinical outcome measurements between the EA and Sham EA groups before and after treatment.

| **Outcome** | **EA group**<br>**(n = 22<sup>#</sup>)** | **Sham EA group**<br>**(n = 18<sup>#</sup>)** | **T value**<br>**(EA vs Sham)** | **P value**<br>**(EA vs Sham)** | **Effect size**<br>**(EA vs Sham)** |
| --- | --- | --- | --- | --- | --- |
| **Number of days with migraine** |  |  |  |  |  |
| Baseline,<br>mean,(sd),(95 % CI) | 7.53(5.42)<br>(4.25 to 10.81) | 6.46(3.46)<br>(4.54 to 8.38) | 0.61 | 0.54 |  |
| End of treatment, mean,<br>(sd),(95 % CI) | 4.92(5.13)<br>(1.81 to 8.02) | 4.57(2.73)<br>(2.99 to 6.15) | 0.21 | 0.82 | 0.086(-0.71 to<br>0.88) |
| Difference in means<br>End of treatment-baseline, mean, (sd),<br>(95 % CI) | 2.61(2.87)<br>(0.87 to 4.35) | 2.07(3.64)<br>(-0.034 to 4.17) | 0.43 | 0.66 | 0.16 |
| t value | 3.28 | 2.12 |  |  |  |
| P value | 0.0065** | 0.053 |  |  |  |
| Effect size | 0.50 (0.17 to 0.82) | 0.65(-0.042to<br>1.35 ) |  |  |  |
| **Frequency of migraines** |  |  |  |  |  |
| Baseline,<br>mean, (sd), (95 % CI) | 4.69(2.05)<br>(3.44 to 5.93) | 4.26(2.28)<br>(3.00 to 5.53) | 0.51 | 0.60 |  |
| End of treatment, mean, (sd), (95 % CI) | 3.15(2.26)<br>(1.78 to 4.52) | 3.57(1.45)<br>(2.73 to 4.41) | -0.56 | 0.57 | 0.22(-0.57 to 1.02 ) |
| End of treatment-baseline, mean, (sd), (95 % CI) | 1.53(2.40)<br>(0.086 to 2.98) | 0.71(2.78)<br>(-0.89 to 2.32) | 0.82 | 0.41 | 0.32 |
| t value | 2.30 | 0.95 |  |  |  |
| P value | 0.039* | 0.35 |  |  |  |
| Effect size | 0.71(7.03e-05 to 1.42 ) | 0.36(-0.44 to 1.17 ) |  |  |  |
| VAS score |  |  |  |  |  |
| Baseline, mean, (sd), (95 % CI) | 4.11(1.32)<br>(3.31 to 4.91) | 4.49(1.60)<br>(3.63 to 5.35) | -0.70 | 0.48 |  |
| End of treatment, mean,(sd), (95 % CI) | 3.24(1.53)<br>(2.31 to 4.17) | 4.29(1.95)<br>(3.24 to 5.33) | -1.60 | 0.12 | 0.585(-0.19 to 1.36 ) |
| End of treatment-baseline, mean, (sd), (95 % CI) | 0.86(1.99)<br>(-0.33 to 2.07) | 0.25(1.74)<br>(-0.71 to 1.22) | 0.85 | 0.40 | 0.33 |
| t value | 1.81 | 0.568 |  |  |  |
| P value | 0.047* | 0.573 |  |  |  |
| Effect size | 0.61 ( -0.26 to 1.47 ) | 0.14 ( -0.36 to 0.64 ) |  |  |  |

\* $P<0.05$ ; \*\* $P<0.01$ . # The baseline characteristics and clinical outcomes were based on the intention-to-treat (ITT) population. We omitted the cases that retained only the baseline measurement but had missing data for all clinical outcomes.

After 4 weeks of EA treatment, patients in the EA group showed a significant reduction in days (*P* = 0.006) and frequency (*P* = 0.039) of migraines as well as a decrease in VAS scores for pain intensity (*P* = 0.047) compared to the baseline period (Table 1). In contrast, patients in the Sham EA group also showed a decrease in all three symptoms, but this decrease was not statistically significant (*P* > 0.05) (Table 1). However, there was no significant difference between the EA group and the Sham EA group in alleviating migraine (Table 1). Our results were consistent with the multicentre trial of acupuncture for migraine by Yin Li (Li *et al*., 2012). Since a significant difference may not explain the real magnitude of the clinical effect of acupuncture, we further calculated the effect size to validate the efficacy of acupuncture in the context of the clinical setting (Zhao *et al*., 2017).

After 4 weeks of treatment, the effect size measured by Cohen’s D in the EA group compared to the baseline period was 0.50 (95% CI, 0.17 to 0.82) for the reduction in the number of migraine days, 0.71 (95% CI, 7.03e-05 to 1.41) for the decrease in the frequency of migraines and 0.61 (95% CI, -0.26 to 1.47) for the decrease in VAS scores (Table 1). In the context of our study, these effect sizes beyond 0.5 in the EA group manifested that the effect sizes in the EA group are, on average, 0.5 standard deviations greater for relieving those symptoms of migraine than those during the baseline period (Ben *et al*., 2020). However, the effect sizes in the Sham EA group compared to the baseline period were 0.65 (95% CI, -0.042 to 1.34) for the reduction in the number of migraine days, 0.36 (95% CI, -0.44 to 1.17) for the decrease in frequency of migraines and 0.14 (95% CI, -0.36 to 0.63) for the decrease in VAS scores (Table 1). These results of effect size were partly in accordance with the large IPD meta-analysis by Vickers et al (Vickers *et al*., 2012), although they were not statistically significant. Specifically, the effect size in the EA group was 0.585 (95% CI, -0.19 to 1.36) compared to the Sham EA group, which means that the EA group is, on average, 0.585 standard deviations greater for decreasing VAS scores than the Sham EA group (Table 1).

### Metabolic discrimination between migraine and healthy controls

We employed ^1^H NMR techniques and Chenomx NMR Suite 4.5 (Chenomx, Calgary, Canada) to identify 22 metabolites that were measured in a total of 50 plasma samples from migraine patients in the baseline period (MA, n = 40) and healthy controls (n = 10) (Figure 2 and Table 2). After excluding outliers of abnormal metabolic profiles and baseline values among included migraine patients using PCA analysis, we first performed orthogonal signal correction for partial least square discriminate analysis (OPLS-DA) on the normalized Carr-Purcell-Meiboom-Gill (CPMG) NMR data to distinguish metabolic profiles of migraine patients in the baseline period from the profiles of the healthy controls. By using SIMICA-P statistical software, OPLS-DA analysis demonstrated a distinct separation between the MA group and healthy controls (Figure 3A x-axis; *R*^2^*Y* = 75.8%). For the above score plot, R^2^Y manifested the proportion of the variance in the y variable explained by the regression model (Wang *et al*., 2014). Thus, the separation in the plot manifested a crucial metabolic phenotype difference between migraine patients and healthy controls. Correspondingly, the loading plot showed the information of the possible metabolites separating the metabolic profiling (Figure 3B), such as glycine, glutamine, and alanine. After we identified the distinct metabolites in the loading plot, we subsequently conducted Lasso regression, ANOVA combined with Box-Cox transformations and Bonferroni correction on the CPMG dataset to further explore potential metabolic biomarkers separating the profiles of migraine patients and healthy controls. The dominant metabolites that discriminate these two groups are presented in Table 2. After Bonferroni correction, we found a significant increase in 5 plasma metabolites, including glycerine (*P* = 0.00085), choline (*P* = 0.004), citrate (*P* = 0.016), pyruvic acid (*P* = 0.049) and glutamine (*P* = 0.0017), and a significant decrease in 3 plasma metabolites, including glycine (*P* = 0.0017), alanine (*P* = 0.002), and lipid (*P* = 0.030) in the MA group compared to healthy controls. We further conducted ROC curve analysis to validate the clinical importance of these biomarkers. The result of the ROC curve analysis showed that citrate performs well in differentiating between migraine patients and healthy persons (area under the curve (AUC) = 0.87, *P* = 0.005) (Figure 4C). Moreover, we conducted OPLS-DA analysis on the ^1^H NMR longitudinal eddy current delay (LED) dataset, and the LED results also illustrated a similar separation between the MA group and the healthy control group (Supplementary Figure 2C and Supplementary Table 2; *R*^2^*Y* = 99.7%). Accordingly, we found a significant decrease in plasma N-acetyl glycoproteins (NAc) (*P*=0.00082) and a significant increase in plasma phosphatidylcholine (Ptdcho) (*P* = 0.035) in the MA group compared to the healthy control group after Bonferroni correction (Table S2). These significantly changed metabolites found in the CPMG and LED results were selected as potential biomarkers for the metabolic features of migraine in the clinical setting.

**Fig. 2:**
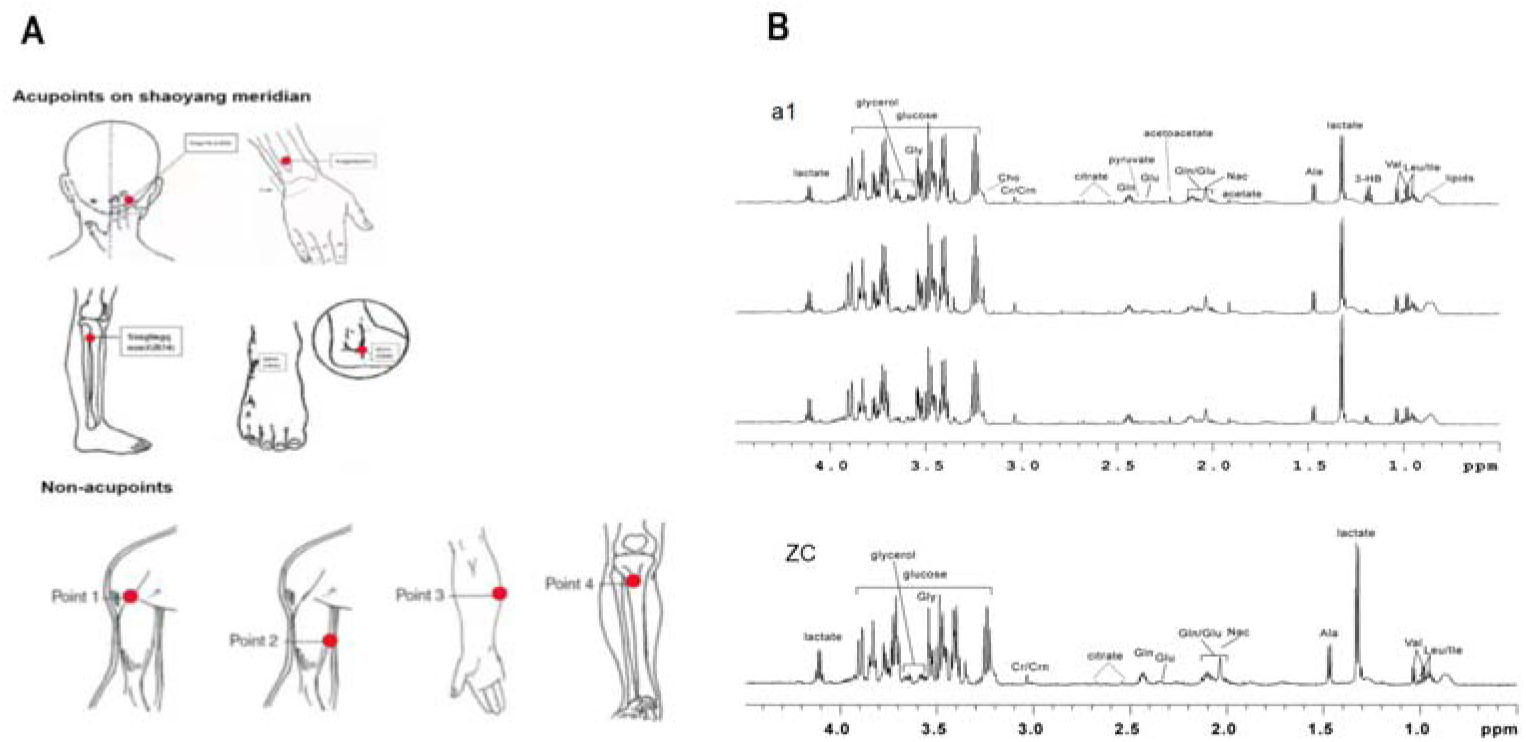
Locations of acupuncture points and typical ^1^H NMR spectra of plasma samples. **(A)** Locations of acupuncture points at Shaoyang meridian and non-acupoints **(B)** Typical ^1^H NMR CPMG spectra of plasma samples. ^1^H NMR experiments were carried out, and Chenomx NMR Suite 4.5 (Chenomx, Calgary, Canada) software was used to identify 22 metabolites measured in a total of 50 plasma samples from 40 migraine patients and 10 healthy controls before and after EA or sham EA treatment (Table 2). a1, migraine patients; ZC, healthy controls; Ala, alanine; Cr/Crn, creatine; Gly, glycine; gln, glutamine; glu, glutamate; Val, valine; 3-HB, 3-hydroxybutyric acid; Leu/lle, isoleucine; Different citrate levels of migraine patients indicate that there is a difference at the citrate level in the spectra.

**Fig. 3.**
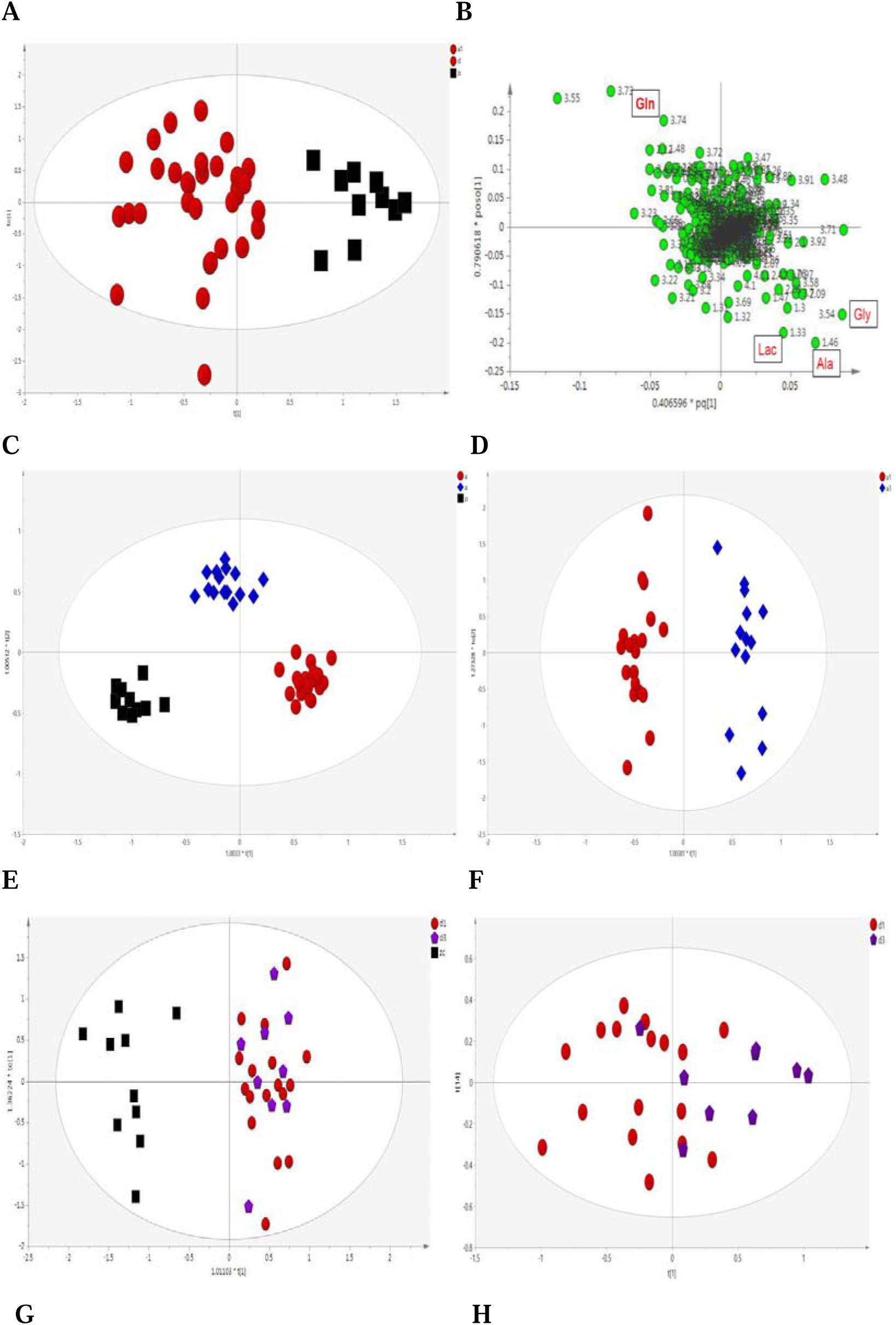
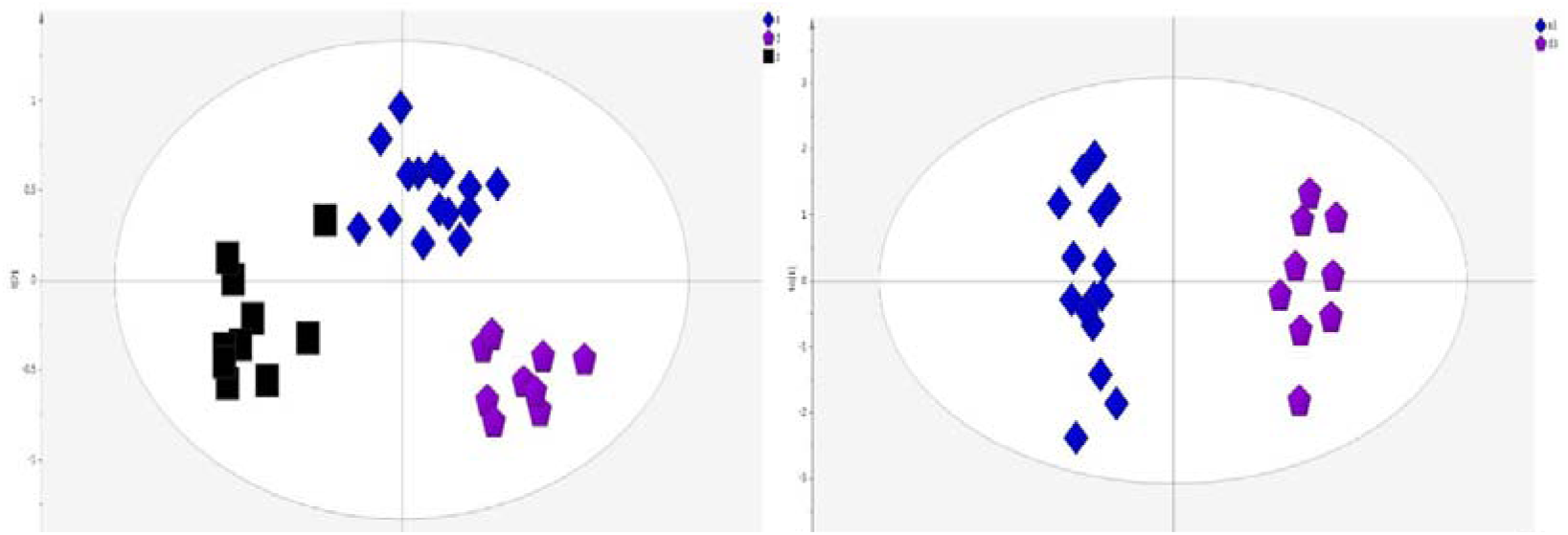
Clear separation of metabolic profiles among groups. OPLS-DA analysis for CPMG data manifested clear separation among migraine patients (red dots), healthy controls (black boxes), migraine patients after 4 weeks of EA treatment (blue diamonds), and migraine patients after 4 weeks of sham EA treatment (purple stars). t[1] and t[2] represent the first and second components in the OPLS-DA result, respectively. The missing samples from the EA group and Sham EA group on the score plots were excluded due to the outlier and drop out. **(A)** Clear separation of metabolic profiling was achieved between migraine (red dots, n=40) and healthy control (black boxes, n=10) groups. **(B)** Corresponding loading plots showing metabolites that may influence the separation for (a). Gly, glycine; Gln, glutamine; Lac, lactic acid; Ala, alanine. **(C)** The separation of metabolic profiling showed that EA treatment (blue diamonds, n=22) reversed the change in metabolic profiling in migraine patients (red dots, n=22) compared with healthy controls (black boxes, n=10) (*Table 3*). **(D)** The results showed a clear discrimination in metabolic profiling between migraine patient after EA treatment (blue diamonds, n=22) and migraine patients before EA treatment (red dots, n=22) (*Table 3*). **(E)** The results showed that migraine patient after sham EA treatment (purple stars, n=18) could not restore the change of metabolic profiling in migraine patient before sham EA treatment (red dots, n=18) compared with healthy controls (black boxes, n=10) (*Table 4*). **(F)** The result showed metabolic profiling of migraine patient before Sham EA treatment (red dots, n=18) could not be discriminated with migraine patient after Sham EA treatment (purple stars, n=10) (*Table 4*) **(G)** The profiling indicated that the metabolic profiling of migraine patients after EA treatment (blue diamonds) was closer to that of healthy controls (black boxes) compared to migraine patients after sham EA treatment (purple stars). **(H)** Clear separation of metabolic profiling was discriminated between EA treatment (blue diamonds) and sham EA treatment (purple stars) (*Table 5*).

**Fig. 4.**
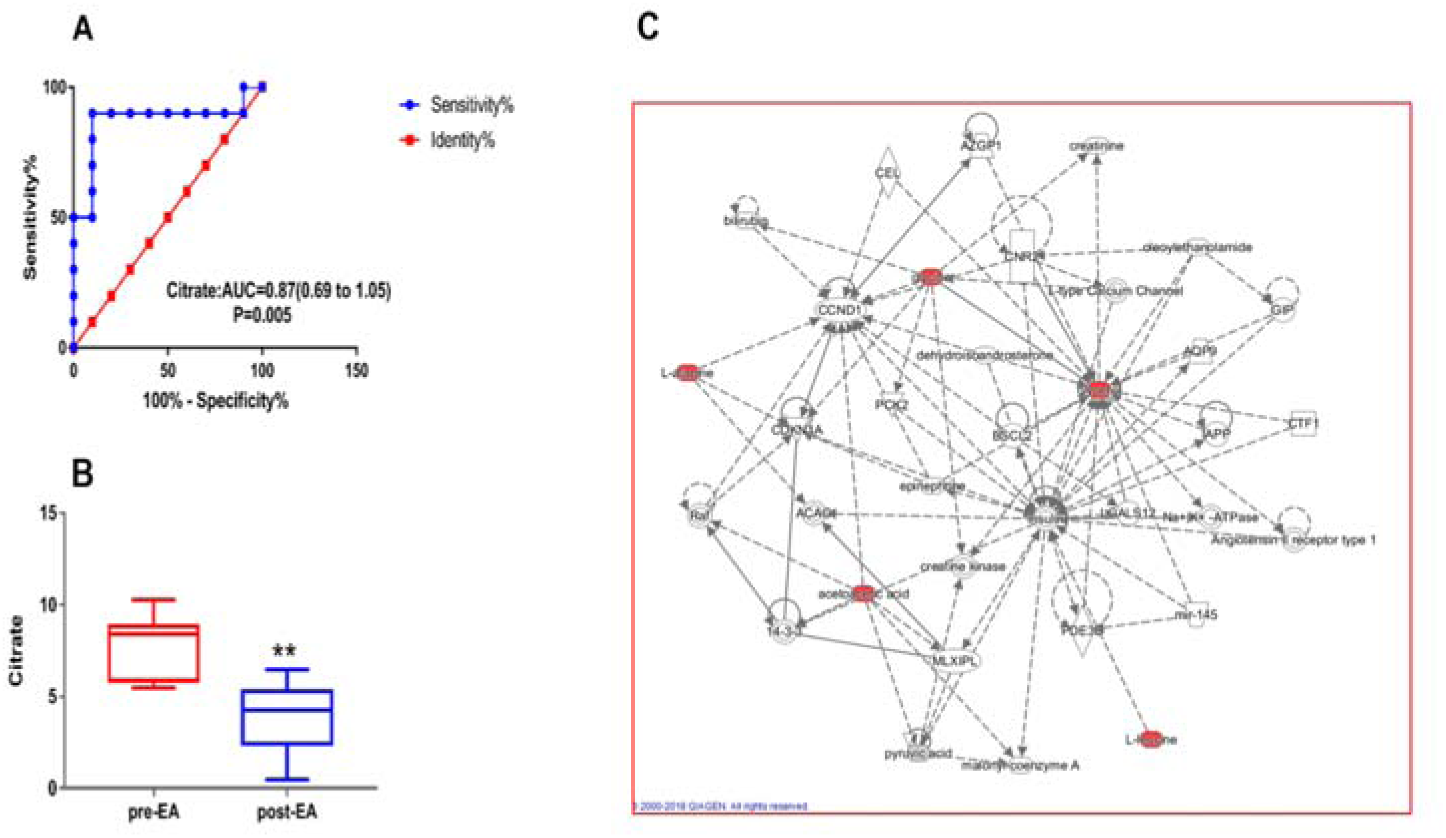
Identification of significant metabolites for the clinical efficacy of acupuncture. We conducted receiver operating characteristic (ROC) curve analysis to validate the significance of potential biomarkers for migraine and acupuncture. **(A)** ROC analysis showed that citrate could significantly discriminate the migraine and control groups and thus might be a potential diagnostic biomarker (AUC=0.87) for migraine diagnosis. **(B)** Citrate was significantly decreased (*P = 0.00079*) after EA treatment. **(C)** Employing IPA network analysis, we found that glycerine (glycerol), glycine, acetone, alanine and leucine might be important metabolites for the metabolic network of EA vs sham EA. Glycerine (glycerol), which is located near the centre of the metabolic network of EA vs sham EA, may be the key metabolite for the efficacy of EA and sham EA (**C)** (*Table 5)*.

**Table 2.**
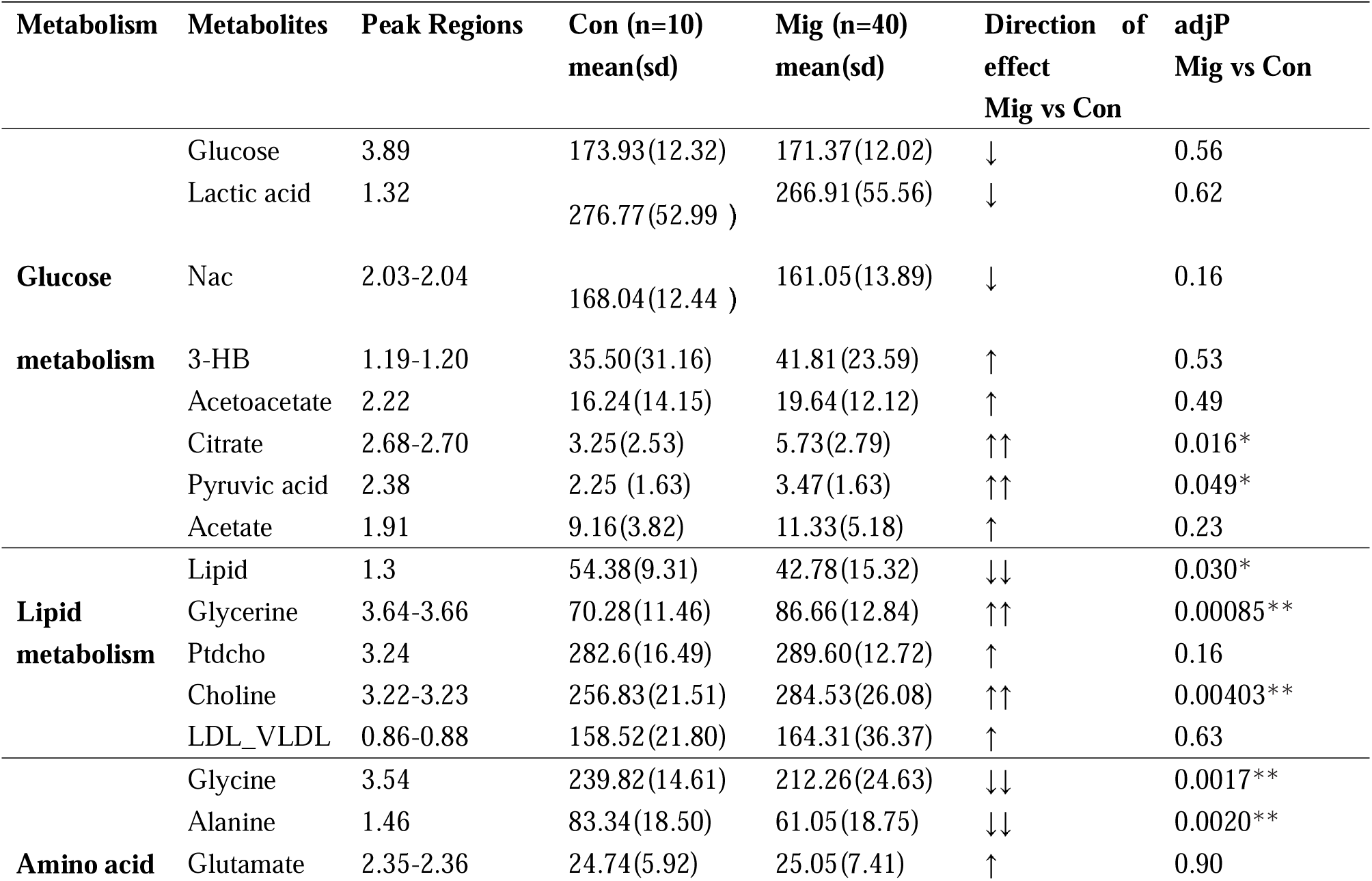
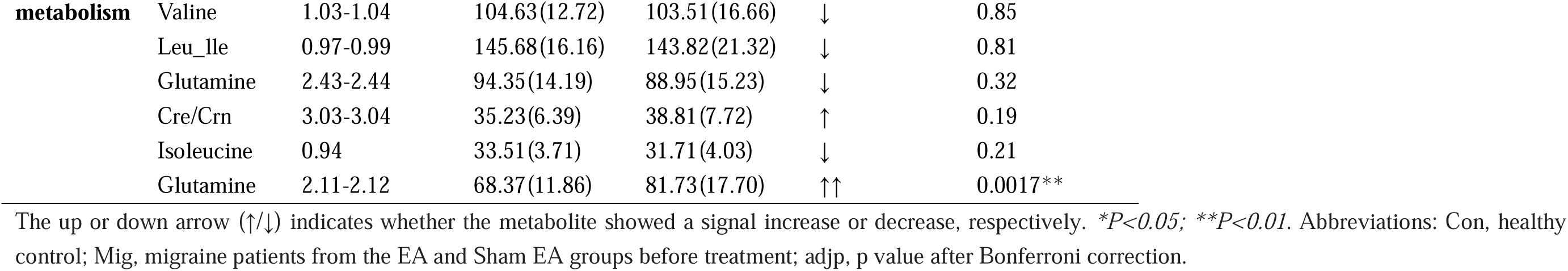
Changes in plasma metabolites in CPMG NMR spectra between healthy controls and migraine patients.

**Table 3.**
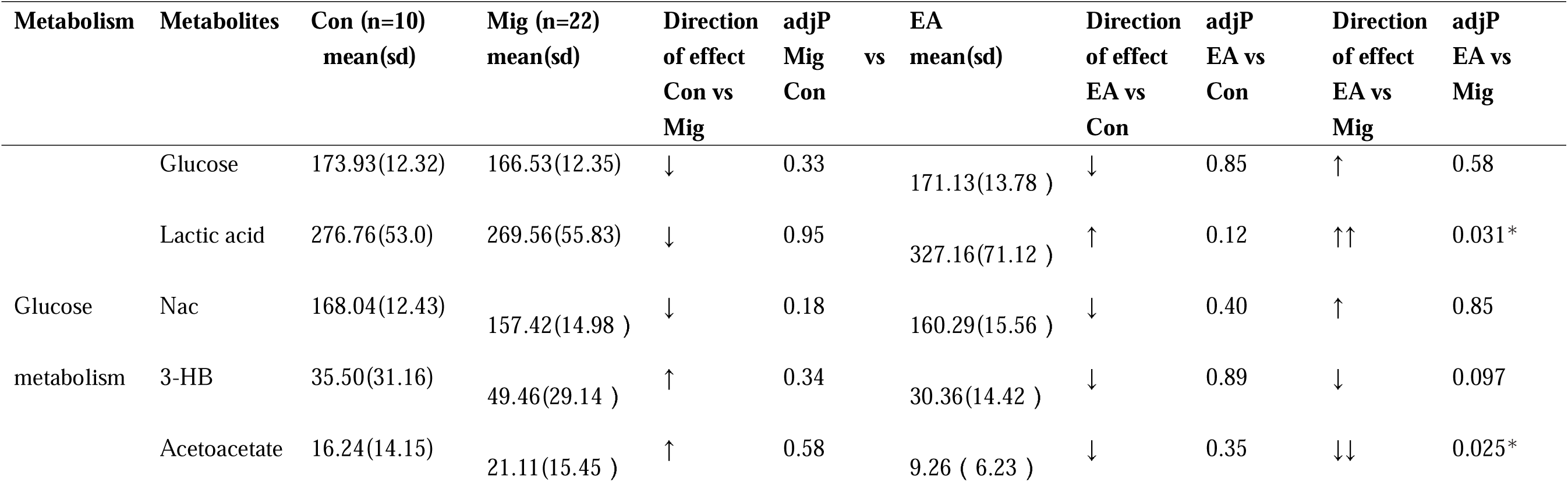
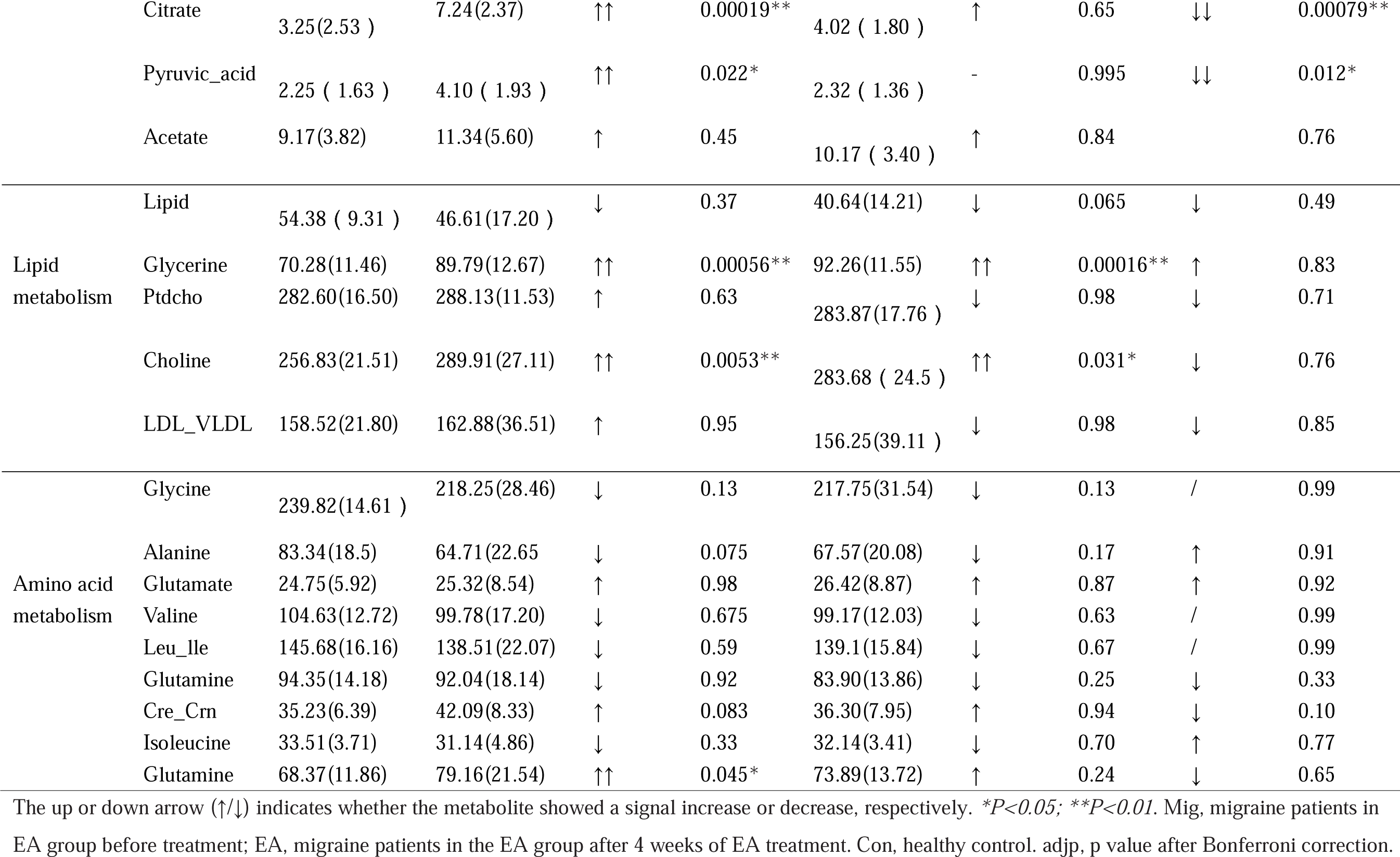
Changes in plasma metabolites in CPMG NMR spectra before and after EA treatment in migraine patients.

**Table 4.**
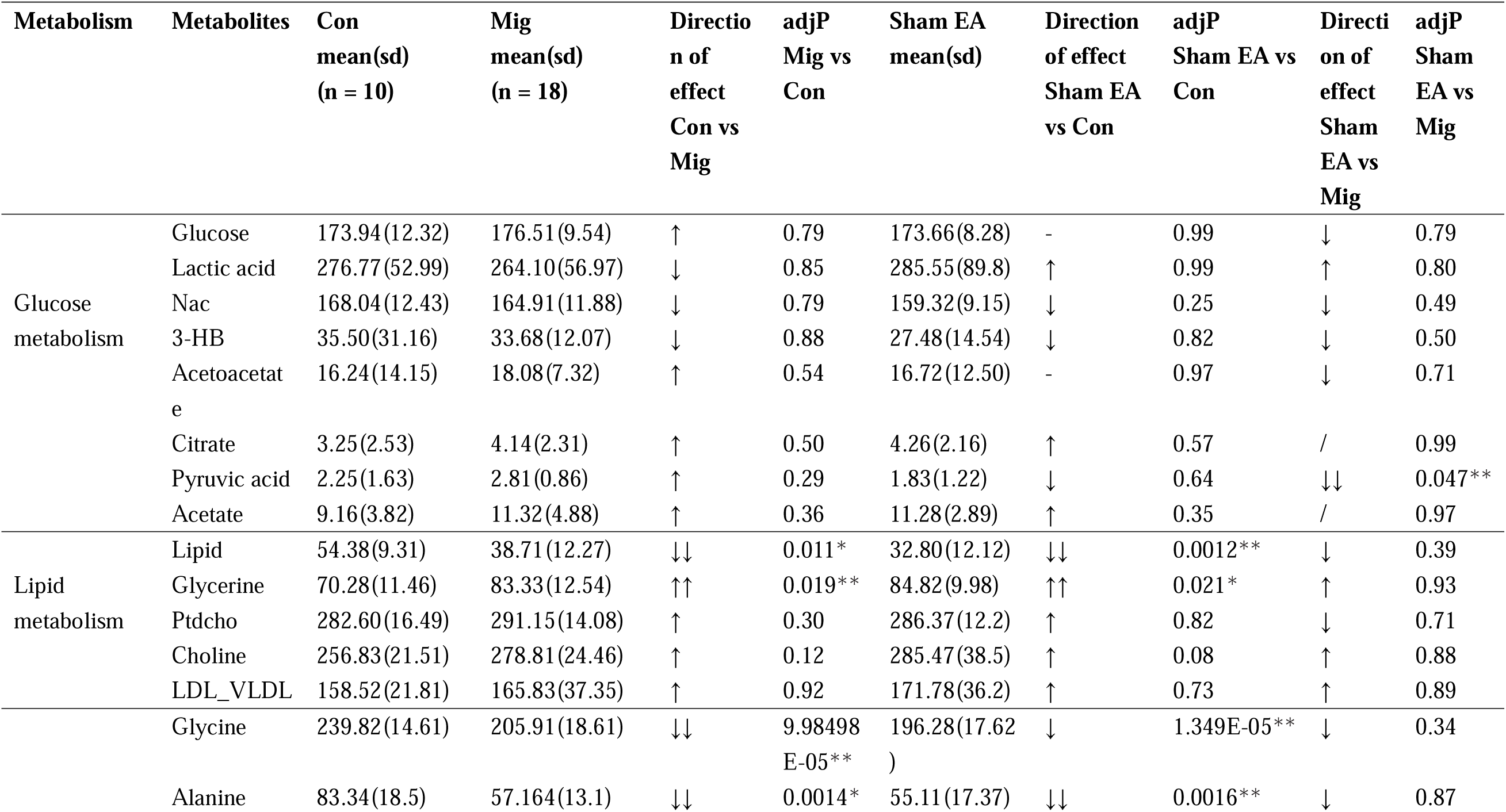
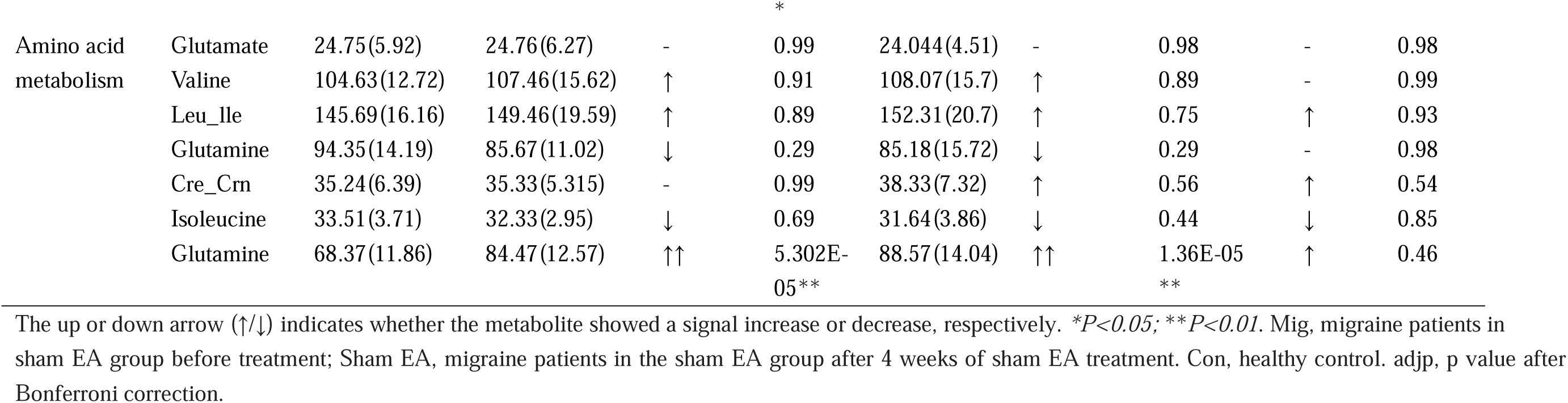
Changes in plasma metabolites in CPMG NMR spectra before and after sham acupuncture treatment in migraine patients.

**Table 5.** Changes in plasma metabolites in CPMG NMR spectra between EA and sham EA treatment.

| Metabolism | Metabolites | EA<br>mean(sd) | Sham EA<br>mean(sd) | Direction of<br>effect<br>EA vs Sham<br>EA | adjP<br>EA vs Sham<br>EA | Coefficient<br>EA vs Sham<br>EA |
| --- | --- | --- | --- | --- | --- | --- |
| Glucose metabolism | Glucose | 171.13(13.78) | 173.67(8.28) | ↑ | 0.64 | n.a. |
|  | Lactic acid | 327.17(71.12) | 285.55(89.88) | ↑ | 0.23 | n.a. |
|  | Nac | 160.29(15.56) | 159.32(9.15) | ↓ | 0.85 | n.a. |
|  | 3-HB | 30.36(14.42) | 27.48(14.54) | ↓ | 0.64 | -0.74 |
|  | Acetoacetate | 9.26(6.23) | 16.72(12.50) | ↑ | 0.061 | 1.06 |
|  | Citrate | 4.02(1.81) | 4.26(2.16) | - | 0.72 | n.a. |
|  | Pyruvic acid | 2.32(1.36) | 1.83(1.22) | ↓ | 0.35 | n.a. |
|  | Acetate | 10.17(3.40) | 11.29(2.90) | ↑ | 0.43 | n.a. |
| Lipid metabolism | Lipid | 40.64(14.22) | 32.80(12.13) | ↓ | 0.18 | n.a. |
|  | Glycerine | 92.26(11.56) | 84.824(9.98) | ↓ | 0.12 | -0.38 |
|  | Ptdcho | 283.87(17.76) | 286.38(12.21) | ↑ | 0.72 | n.a. |
|  | Choline | 283.68(24.50) | 285.47(38.52) | ↑ | 0.87 | n.a. |
|  | LDL_VLDL | 156.25(39.12) | 171.78(36.19) | ↑ | 0.35 | 0.24 |
| Amino acid metabolism | Glycine | 217.75(31.53) | 196.29(17.62) | ↓ | 0.076 | -0.179 |
|  | Alanine | 67.57(20.08) | 55.12(17.37) | ↓ | 0.14 | -0.32 |
|  | Glutamate | 26.42(8.87) | 24.04(4.51) | ↓ | 0.43 | n.a. |
|  | Valine | 99.17(12.03) | 108.07(15.70) | ↑ | 0.13 | n.a. |
|  | Leu_Ile | 139.10(15.84) | 152.31(20.75) | ↑ | 0.088 | 0.19 |
|  | Glutamine | 83.90(13.86) | 85.18(15.72) | ↑ | 0.83 | n.a. |
|  | Cre_Crn | 36.30(7.95) | 38.34(7.32) | ↑ | 0.55 | n.a. |
|  | Isoleucine | 32.14(3.41) | 31.64(3.86) | ↓ | 0.75 | n.a. |
|  | Glutamine | 73.89(13.72) | 88.56(14.04) | ↑↑ | 0.0098** | 0.23 |

The up or down arrow (↑/↓) indicates whether the metabolite showed a signal increase or decrease, respectively. \* $P < 0.05$ ; \*\* $P < 0.01$ .
The coefficient was calculated by Lasso regression. EA, migraine patients in the EA group after 4 weeks of EA treatment; Sham EA, migraine patients in the Sham EA group after 4 weeks of Sham EA treatment.

### Acupuncture reversing metabolic profiling and plasma citrate of migraine

The main objective of our study was to investigate the possible metabolic mechanism for the effectiveness of acupuncture treatment for migraine. Thus, after demonstrating the metabolic profiling of migraine patients, we next explored whether acupuncture changed the metabolites of migraine patients after 20 sessions of EA treatment. Similarly, we performed OPLS-DA analysis on metabolomic data between migraine patients during the baseline period (MA; n = 22) and migraine patients after acupuncture treatment (EA; n = 22). To determine the trend of acupuncture altering metabolites, we also conducted an OPLS-DA analysis on the metabolomic data of the two aforementioned groups and 10 healthy controls (n = 10). After 20 sessions of EA at Shaoyang Meridian acupoints over 4 weeks, we observed that the metabolic profiles of the EA group were close to those of the healthy control group but differed from those of the group at baseline (Figure 3C; *R*^2^*Y* = 94.5%), showing that EA restores the metabolic profiles of migraine similar to those of healthy controls. We further achieved a clear discrimination in metabolic profiling between migraine patients during the baseline period and migraine patients after acupuncture treatment (Figure 3D; *R*^2^*Y* = 85.2%). Notably, we demonstrated a significant decrease in the plasma levels of citrate (*P* = 0.00079) and pyruvic acid (*P* = 0.012) in the EA group compared to the MA group after Bonferroni correction (Table 3). This result indicated that EA relieves migraine by altering the levels of plasma citrate and pyruvic acid in the clinical setting. In addition, we found a significant increase in plasma lactic acid (*P* = 0.031) and a significant decrease in plasma acetoacetate (*P* = 0.025) in the EA group relative to the group at baseline before EA treatment. Further, we found that glycerine (*P* = 0.00016) was significantly elevated in the EA group compared to the healthy control (Table 3). These significantly changed metabolites were potential metabolite biomarkers for EA relieving migraine. The LED data also showed a clear differentiation between migraine patients during the baseline period and migraine patients after acupuncture treatment (Figure S2C; *R*^2^*Y* = 99%). A significant decrease in NAC (*P* = 0.00073) was found in the EA group compared with the healthy control group after Bonferroni correction (Table S3). Taken together, these data demonstrated that EA could enhance anaerobic glycolysis by lowering the plasma levels of citrate and pyruvic acid as well as increasing the plasma level of lactic acid. These regulations in energy metabolism and the tricarboxylic acid cycle (TCA) cycle may be the basis of how acupuncture restores metabolic profiles and relieves migraine in a clinical setting.

### Metabolic basis for the specific effect of acupuncture

The second objective of our study was to detect the metabolic basis for the specific effect of acupuncture in comparison with sham acupuncture. To address this question, we next performed an OPLS-DA analysis on both CPMG and LED data comparing true acupuncture (EA, n = 22) and sham acupuncture (Sham EA, n = 18) groups. To distinguish the trend of acupuncture adjusting metabolic profiles, OPLS-DA analysis was performed on metabolomic data from healthy controls (n = 10), true acupuncture (EA, n = 22), and sham acupuncture (Sham EA, n = 18) groups. After 20 sessions of Sham EA treatment on non-acupoints for 4 weeks, we found that the metabolic profile of the Sham EA group (patients in the Sham EA group after treatment; n = 18) was separated from the healthy controls but similar to the MA group (patients in the Sham EA group before treatment; n = 18) (Figure 3E, *R*^2^*Y* = 48.9%). Moreover, the OPLS-DA analysis was unable to differentiate MA group and Sham EA group (Figure 3F, *R*^2^*X* = 89.9%). In addition, the OPLS-DA results also demonstrated an important discrimination in metabolic profiling among EA, Sham EA and healthy controls (Figure 3G; *R*^2^*Y* = 88.5%), which illustrated that the metabolic profiling of the EA group was close to the healthy controls in relation to the Sham EA group. The difference in the metabolic profiles suggested that EA treatment, not Sham EA treatment, could reverse the metabolic profiles of migraine patients to healthy controls. Further, the OPLS-DA results showed a clear separation of the metabolic profiles between the EA group and Sham EA group (Figure 3H, *R*^2^*Y* = 94.8%), which indicates a discriminated metabolic phenotype between acupuncture and sham acupuncture. The cross-validation for the OPLS-DA analysis of the aforementioned groups was conducted using CV-ANOVA within SIMICA-P software, and the results are provided in Supplementary Table S6.

Corresponding to the OPLS-DA analysis, after Bonferroni correction, ANOVA results showed a significant decrease in pyruvic acid (*P* = 0.047) in the Sham EA group compared to the MA group (Table 4). Similarly, we found a significant decrease in lipid (*P* = 0.0012), glycine (*P* = 0.000013), and alanine (*P* = 0.0016) as well as a significant increase in glycerine (*P* = 0.021) and glutamine (*P* = 0.000013) in the Sham EA group compared with the healthy control (Table 4). These metabolite changes induced by Sham EA reflect the potential metabolic basis for the effect of sham acupuncture on migraine. Specifically, by using Lasso regression analysis, glycine, glycerine, alanine, 3-hydroxybutyric acid (3-HB), isoleucine (leu_lle), low density lipoprotein_very low density lipoprotein (LDL_VLDL), acetoacetate and glutamine were further selected as potential biomarkers for the discrimination of EA and Sham EA (Table 5). Notably, we found that glutamine (*P* = 0.0098) was significantly decreased in the EA group compared to the Sham EA group (Table 5). Combined, these data demonstrate that there is a distinct metabolic difference between EA and Sham EA. These metabolic differences, which can be caused by plasma changes in glycine, glycerine, alanine, 3-HB, leu_lle, LDL_VLDL, acetoacetate, and glutamine, are a potential metabolic basis for the specific effects of acupuncture.

### Functional regulation and network adjustment by acupuncture

After pinpointing the potential metabolic biomarkers for the clinical efficacy of acupuncture, we conducted a metabolic pathway and network analysis among the healthy control, MA, EA and Sham EA groups to further explore the potential upstream pathological mechanisms of acupuncture for migraine. By using IPA, we observed a significant change in the tRNA charging pathway (*P* = 0.031) and a top-changed metabolic network of carbohydrate metabolism, molecular transport, and small molecule biochemistry (score = 14) (Krämer *et al*., 2014) between the healthy control and MA groups (Figure S3A, D).

After 20 sessions of EA treatment for 4 weeks, we found that the pyruvate fermentation to lactate pathway (*P* = 0.0175) was significantly reversed between the MA and EA groups and that the carbohydrate metabolism, energy production, and lipid metabolism networks (score = 9) were the top changed networks between the MA and EA groups (Figure S3B, E). These results manifested that acupuncture reversed the levels of citrate and pyruvic acid through pyruvate fermentation to the lactate pathway, and changed the similar carbohydrate metabolism network, which was altered in migraine patients compared to healthy controls. The changed network of energy production in the EA group further supports the effect of EA based on regulations in energy metabolism from our previous results. Specifically, we found that the tRNA charging pathway (*P* = 0.0307) was significantly discriminated between the EA group and Sham EA group (Figure S3C), and the top network discriminating the EA group and Sham EA group was carbohydrate metabolism, molecular transport, and small molecule biochemistry (score =14, Figure 4C). Glycerine was located near the center of the metabolic network of EA compared to Sham EA (Figure 4C). These findings indicate that the metabolic difference found between EA and Sham EA treatment was linked to the tRNA charging pathway and carbohydrate metabolism, molecular transport, and small molecule biochemistry. These significantly changed metabolic pathways and networks may be the functional basis of the specific effects of EA compared with sham EA.

## Discussion

Here, we have described the first metabolomic study of acupuncture for migraine derived from a randomized controlled trial. By using validated NMR techniques and well-designed machine learning strategies, we demonstrated that EA at acupoints could restore energy deficiency and adjust plasma citrate levels, thereby alleviating migraine. Using advanced Lasso regression models, we first determined 8 metabolic biomarkers that can be attributed to the potential metabolic basis for specific effects of acupuncture compared to sham acupuncture. Taken together, these results demonstrated a distinct metabolic phenotype that can serve as a novel scientific explanation for the efficacy of acupuncture in the clinical setting.

The strength of this study includes a concealed central randomization and strict machine learning statistical strategy. Compared with other metabolomics studies for acupuncture, the use of central randomization in our study could minimize the selection bias, and the independent of statistician could also lower the detection bias, thus improve the reliability and validity of our study result in clinical setting (Zhao *et al*.,2017). Specially, Lasso regression was executed, to eliminate the multicollinearity between metabolites and the inter-metabolite relationships comprising the metabolic network (LeWitt *et al*.,2017, Menni *et al*., 2017). By using this shrinkage method, 8 potential biomarkers distinguishing the acupuncture and sham acupuncture can be selected. Moreover, Box-cox transformation was applied before the ANOVA analysis, which could be a powerful procedure for adjusting skewed distributions and continuous variables, thereby improving accuracy of statistical results (Yu *et al.,*2022). Further, Bonferroni correction, one of the strictest multiple testing correction methods, was executed in the study. This method not only could control the false positive rate, but also pinpointed the validated biomarker for acupuncture (Sedgwick *et al*.,2014). Simply put, we presented a novel and practical machine learning strategy in a randomized acupuncture trial, which could set a possible example for handling high-dimensional metabolomics data in acupuncture trials.

In this study, we found that EA was more clinically important to patients than sham EA, especially for relieving the intensity of pain in migraine patients. First, EA, not sham EA, could significantly alleviate these clinical symptoms in migraine. Second, the effect size of the VAS score for pain relief in the EA group was markedly higher than that in the Sham EA group (Table 1). The EA group showed an average decrease of 1.05 cm more in the total 10 cm VAS compared with the Sham EA group, and this decrease reached the 1 cm MID in the VAS scale (Thorlund K *et al*., 2011), which accounted for 10.5% greater pain relief than Sham EA treatment at clinic. These results were further in agreement with a long-term randomized trial on acupuncture for migraines (Zhao *et al*.,2017). In addition, the nonsignificant results observed in both acupuncture groups after treatment can be attributed to the limit of statistical significance for small sample sizes as well as the penetration needles in sham EA, which had already been analyzed in a large IPD meta-analysis (Vickers *et al*., 2012; Li *et al*., 2012).

Recently, accumulating evidence has confirmed that energy deficiency and mitochondrial dysfunction are two cornerstones of migraine pathophysiology (Lodi *et al*.,2006; Sparaco *et al*.,2006). A novel metabolic picture of migraine has been presented in a publication of Nature Reviews Neurology (Gross *et al*.,2019): Decreased cerebral glycogen prolonged synaptic activity, increased cerebral excitability, reduced CSD threshold, and thus stimulated CSD in migraine. On the other hand, mitochondrial dysfunction induced excessive production of free radicals and subsequently activated transient receptor potential (TRP) channels that increase CGRP release, which is pivotal for mediating migraine attack. Compared with former studies, we have shown enhanced aerobic glycolysis and reduced gluconeogenesis, which trigger the increase in citrate as a key intermediate in the TCA cycle and lead to mitochondrial energy metabolism disorder in migraine patients. Lipid metabolism is also affected and disordered, accompanied by an increase in fat mobilization, glycerine and ketone bodies (Table 2, Figure 5) (Aurora *et al*.,2007; Bélanger *et al*., 2011). These findings further detailed the imbalance between reduced energy supplies and increased energy needs among migraine patients (Figure 5). In addition, we demonstrated that citrate performed well in discriminating migraine patients and healthy controls, revealing a potential novel biomarker for migraine. It has been shown that an increase in citrate could activate transient receptor potential channel A1 (TRPV1) pain receptors, enhance the protein kinase B (AKT) signaling pathway (Figure S3D) (Gross *et al*., 2019; Liu et al., 2017; Xu *et al*., 2018; Chen *et al*., 2017), thereby trigger migraine (Table 2, Figure 5). Collectively, our findings not only expanded the pathophysiology of migraines with an additional understanding of metabolism, but also highlighted the demands of efficient metabolic strategies for migraine treatment.

**Fig. 5.**
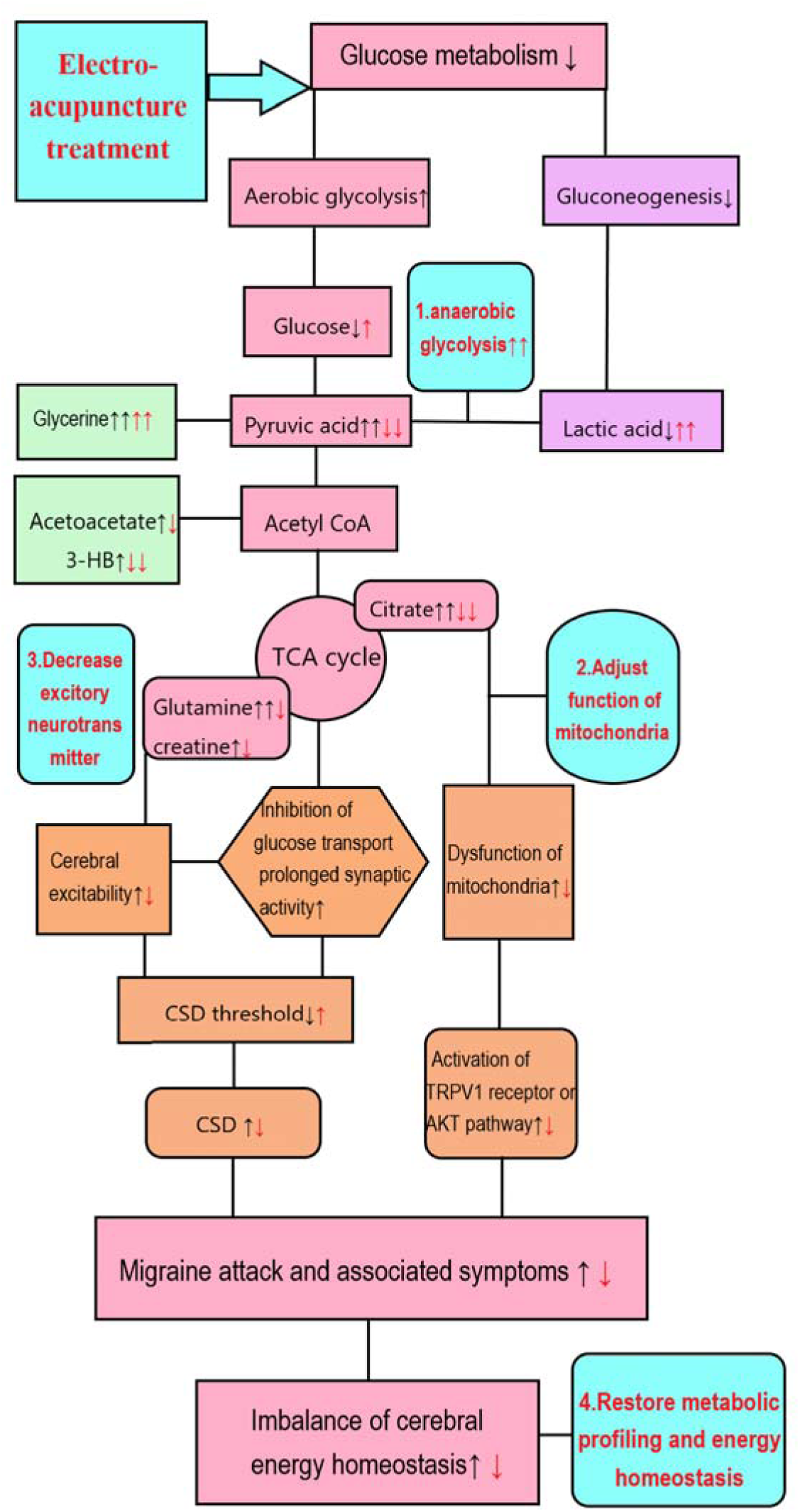
The metabolic mechanism for the efficacy of acupuncture in migraine. The black arrow (↑/↓) shows the depletion of glucose metabolism and increased lipid metabolism in migraine patients, leading to energy deficiency and disorder of the TCA cycle and mitochondria, which trigger migraine attack. The red arrow (↑/↓) and the blue box indicate the possible metabolic mechanism of acupuncture, which suggests that EA may restore energy deficiency by enhancing anaerobic glycolysis and lowering plasma citrate levels in the TCA cycle, thus decreasing migraine attack and restoring mitochondrial function and metabolic profiling in migraine patients.

Previously, there is little understanding about the metabolic basis of acupuncture in relieving migraine. Here, we provided novel metabolic evidence of EA relieving migraine as follows: (i) EA could enhance anaerobic glycolysis through converting increased pyruvic acid into lactic acid and subsequently decrease the elevated level of citrate in the TCA cycle and acetoacetate in lipid metabolism in migraine patients (Figure 5). (ii) Importantly, citrate, which was found to be significantly increased in migraine patients, was significantly decreased after EA treatment, which provides direct metabolic evidence of EA adjusting mitochondrial function in migraine (Table 2, Figure 5). To date, both experimental and clinical evidence has shown that the depletion of glycogen and hypoglycaemia can trigger migraine attacks (Lodi R *et al*., 2006; Pearce *et al*., 1971; Hockaday *et al*., 1971). And lactic acid is a prior energy source for neurons during brain energy deficiency (Suzuki *et al*., 2011). Therefore, the first possible metabolic mechanism for EA relieving migraine is the increased anaerobic glycolysis caused by EA combined with the increase in lactic acid, which can quickly compensate for the lack of energy and thereby possibly reduce the CSD triggering migraines (Figure 5). Accordingly, a PET-CT study (Zeng *et al*.,2012; Yang *et al*.,2012) also revealed that EA could restore glucose metabolism in key regions of the descending pain modulation system (DPMS) of brain for migraine. Interestingly, our findings identified a potential subgroup of migraine patients with a distinct metabolic phenotype that could benefit from EA treatment in clinics. Moreover, previous results have indicated an important role of mitochondrial dysfunction in migraine and the positive association between citrate levels and activation of the AKT signaling pathway (Gross *et al*., 2019; Liu et al., 2017; Xu *et al*., 2018; Chen *et al*., 2017). Recent animal experiment further demonstrated inhibition of the activation of AKT could attenuate cumulative pain score and pain-related behaviors (Xu *et al*., 2019). Thus, the second potential metabolic evidence of EA managing migraine is that EA could adjust mitochondrial dysfunction and inhibit the pain-related AKT pathway by decreasing the plasma level of citrate, thereby alleviating pain in migraine (Figure 5). In particular, this metabolic biomarker of citrate may serve as the desired quantitative biomarker for further clinical acupuncture research. Third, in the previous decades, profound acupuncture experiments focused on the pivotal role of opioid peptides and its related Arc-PAG-NRM-spinal dorsal horn pathway in mediating acupuncture analgesia (Zhao *et al*.,2008; Huang *et al*.,2002). Compared to those neurological evidence of acupuncture analgesia, our results revealed a systemic modulation effect of EA on both energy metabolism and mitochondrial function in migraine patients. These findings opened exceptional new insights into the metabolic mechanism underlying the effectiveness of acupuncture analgesia and could be developed into potent non-opioid drugs and multi-metabolic therapeutic targets for future migraine treatment.

The specific effect of acupuncture is another long-debated issue among clinical acupuncture trials. Fei Y et al and Prof. Gordon Guyatt pointed out in the publications of BMJ that one of the specific methodology challenges in acupuncture trials is the estimation for the optimal treatment effect of acupuncture compare to sham acupuncture (Fei *et al*., 2022). Here, we presented a unique metabolic mechanism for the specific effect of acupuncture: We demonstrated that there are important but different ways of energy supply between EA and Sham EA for relieving migraine. (i) EA specifically reversed the deficiency of energy metabolism in migraine patients through anaerobic glycolysis compared with Sham EA. In detail, EA can restore energy deficiency by enhancing anaerobic glycolysis and decomposition of acetoacetate (Table 2, Figure 5) in the plasma (Salek *et al*.,2017; Peek *et al*.,2020), which could quickly compensate for the lack of energy and thereby reduce the CSD triggering migraine. In contrast, sham EA may partially supply energy to migraine patients by fat mobilization through lipid metabolism (Table 3). In addition, the network analysis showed that glycerine, a classic product of fat mobilization, was the key node in the metabolic network of EA compared with sham EA (Figure 4C). Therefore, we can postulate the hypothesis that these different ways of supplying energy may be one of the crucial factors contributing to the different effect between acupuncture and sham acupuncture. (ii) EA specifically relieves migraine by reversing citrate, thereby adjusting mitochondrial dysfunction compared with sham EA. The TCA cycle is the centre of energy metabolism in the body. In particular, we have shown that EA, not sham EA, could reverse plasma citrate and subsequently adjust the dysfunction of the TCA cycle in mitochondria, thus restoring metabolic profiling and relieving migraine in migraine patients (Figure 5). (iii) We found that glutamine, a classic excitatory neurotransmitter that facilitates CSD and is linked to the intensity of migraine (Gao *et al*.,2014; Aroke *et al*.,2020; Alam *et al*.,1998), showed a Bonferroni-corrected significant decrease in the EA group compared with the Sham EA group. This result is consistent with our previous metabolomic study of an acute migraine rat model (Gao *et al*.,2014). The significant decrease in glutamine in the EA group relative to the Sham EA group further explains why EA had a better clinical migraine pain relief effect than sham EA. In addition, our study found that both EA and sham EA could significantly increase glycerine levels, thereby supplying energy to migraine patients. This finding gives a possible explanation for the non-specific effect of EA and sham EA observed in these clinical trials. Collectively, the specific effect of EA on both anaerobic glycolysis and mitochondrial function for migraine exclusively provides a possible scientific mechanism of the efficacy of acupuncture for clinical doctors and clinical decision-makings.

In summary, our findings indicate that EA specifically relieves migraine by enhancing anaerobic glycolysis and decreasing the plasma levels of citrate and glutamine among migraine patients. Our study is of great importance because it provides scientific evidence and explanations for the specific effect of acupuncture relieving migraine for both clinical doctors and policymakers. By applying modern machine learning techniques, this metabolic evidence could enlighten a brand new direction into acupuncture analgesia mechanism, which in turn would pose fresh challenges for future acupuncture researches.

## Limitations of the study

The main limitation of this study is the limited sample size and the non-targeted metabolic technique which cannot detect targeted metabolite changes in the TCA cycle and glucose metabolism. And the validated animal experiment should also be executed. Future studies will extend this work using a larger sample size of migraine patients and targeted metabolomics on both human and animals for possible validation experiments.

## Materials and methods

### Ethics approval

The clinical trial protocol was approved by the Ethics Review Committee of the 1st Teaching Hospital of Chengdu University of TCM and had been published [2007KL-002] (Li *et al*., 2008). The multicentre randomized trial was registered (clinicaltrials.gov: NCT00599586). All study procedures were designed and conducted in accordance with principles of the Declaration of Helsinki and the Chinese version of the International Conference on Harmonisation --- Good Clinical Practice. All patients signed written informed consent.

### Participants

From April 1, 2008, to August 12, 2009, 476 migraine patients were recruited in the multicentre trial. And the migraine patients included in this metabolomic study were all recruited from a single trial centre, the 1st Teaching Hospital of Chengdu University of TCM, which is one of the clinical centers in the multicentre trial. To be included, the migraine patients had to meet the International Headache Classification criteria for migraines (ICHD-II 2004) (Headache Classification Subcommittee of the International Headache Society, 2004), as diagnosed by a doctor. The inclusion criteria were as follows: age from 20 to 45 years old; female only; Han nationality; onset of migraines before age 45; more than 1 year of migraine history and acute migraine attacks 2 to 8 times per month during the previous three months; no use of any prophylactic drug for migraine during the previous 2 weeks; body mass index (BMI) ranging from 18-24; no heart, liver, or kidney disease detected by ultrasound or blood tests as well as other serious organic diseases; willingness to complete 20 acupuncture treatments for 4 weeks; and ability to provide written informed consent.

We excluded patients who had headache owing to organic disorders (e.g., cerebrovascular disease, vascular malformation, arthritic conditions, arteriosclerosis or hypertension) or patients with psychosis, pregnancy or lactation, allergies, bleeding disorders or serious diseases of the heart, liver, kidney or other organs.

Healthy females without migraine who had no significant differences in age, gender, or BMI compared to migraine patients were recruited as the healthy control group.

### Randomization and interventions

The overall trial approach is shown in Figure 1. Forty female patients who met the inclusion criteria were randomly assigned to the electroacupuncture group at specific acupoints belonging to the Shaoyang Meridian group (EA) or the electroacupuncture group at non-acupoints (Sham EA) using a central randomization procedure controlled by the Chengdu Good Clinical Practice (GCP) Center. Central randomization was conducted by a GCP center computer. The independent study assistant sent the patient’s information to the GCP centre through email or short message service (SMS) message. Random numbers and group assignments were accordingly generated by a GCP computer and sent back to the study assistant by a feedback email or SMS. Acupuncturists were not involved in patient recruitment in this trial. Patients and outcome assessors were blinded to randomization. Patients were informed that they would receive one of two types of acupuncture treatment: one was based on traditional Chinese acupuncture theories and another was using modern acupuncture theory.

Acupuncture treatment was performed unilaterally, alternately at specific acupoints of Waiguan (TE5), Yanglingquan (GB34), Qiuxu (GB40), and Fengchi (GB20), which belong to Shaoyang Meridian of the hand and foot in the EA group (Figure 2A). Deqi sensation was required for the EA group. In contrast, sham acupuncture treatment was conducted unilaterally, alternately at predefined non-acupoints, including the middle point between the tip of the elbow and the axilla, the middle point between the epicondylus medialis of the humerus and the ulnar side of the wrist on the ulnar side, the edge of the tibia 1 to 2 cm lateral to the Zusanli (ST36), and the anterior border of the insertion of the deltoid muscle at the junction of the deltoid and biceps muscles in the medial arm (Li *et al*.,2008) (Figure 2A). Deqi sensation was not required for the Sham EA group. All acupoints and sham acupoints were punctured by disposable stainless steel needles (0.25LJmmLJ×LJ40LJmm; 0.25LJmm ×LJ25LJmm; Suzhou Hwato Medical Appliance Co., Ltd., 2270202, Suzhou City, China) at a depth of 20 mm-40 mm. In addition to normal puncturing, 4 auxiliary needles were punctured 2 mm beside every acupoint or non-acupoint with a depth of 2 mm without manual stimulation (Li *et al*.,2008). Transcutaneous electroacupuncture stimulation was subsequently conducted at the acupoints or sham acupoints for 30 minutes using a Han’s acupoint nerve stimulator (HANS-200, Nanjing, China) after needle insertion. The stimulation frequency was set at 2/15 Hz, and the intensity varied from 0.1 mA to 1 mA, adjusted in accordance with the patient’s perception. All acupuncture procedures were conducted in accordance with standards for reporting interventions in clinical trials of acupuncture (STRICA) (MacPherson *et al*., 2010) and a predefined acupuncture SOP by a licensed acupuncturist with at least 5 years of clinical acupuncture experience and with a completed postgraduate degree. Acupuncture treatment was performed 5 times per week according to the patient’s convenience, and a total of 20 acupuncture treatments were performed on both the EA and Sham EA groups. The patients were instructed not to take regular medications during acupuncture treatment. However, ibuprofen (300 mg each capsule with sustained release) could be used as rescue medication when severe migraine attacks happened.

### Outcomes and sample collection

The number of days with migraine before and after four weeks of treatment was defined as the primary outcome for evaluating the frequency of migraine attack. This outcome was recorded in a migraine diary by the migraine patients during the baseline and four-week treatment period. Together with the number of days with migraine, the frequency of migraine and the visual analog scale (VAS) were also recorded by the migraine patients in the same period. The visual analog scale (VAS) is a validated and reliable instrument for assessing pain severity and alleviation. On this scale, pain is measured by placing a handwritten mark on a 10-centimeter line representing a continuum between “no pain” (0 cm) and “worst pain” (10 cm) (10cm). This provides a pain severity rating in centimeters out of ten, such as six out of ten (or 6/10) (Delgado *et al*., 2018). Measurements of the frequency and pain intensity of migraine were secondary outcomes. A total of two blood samples were collected for metabolomics analysis at the start of acupuncture treatment and at the end of 4 weeks of treatment. All fasting venous blood samples (approximately 5 mL) were collected at approximately 7:30–9:30 am and then stored at -80°C.

### NMR experiments

^1^H NMR spectra of the plasma samples were collected and analysed as described in our previous study (Gao *et al*., 2014). 1 sample from acupuncture group (migraine patients after acupuncture treatment) and 1 sample from sham acupuncture group (migraine patients after sham acupuncture treatment) were not qualified to be analyzed before experiment. Prior to NMR analysis, plasma samples were thawed and centrifuged at 13000 × g, 4 *^∘^*C for 10min. 300 *µ*L of each supernatant was transferred into 5mm NMR tube, mixed with 250 *µ*L of D_2_O for field frequency lock and 50 *µ*L of 3-trimethylsilyl- ^2^H_4_-propionic acid sodium salt (TSP) in D_2_O (1mg/mL) as chemical shift reference. All samples contained a final volume of 600 *µ*L and were vortexed repeatedly.

^1^H NMR data of plasma were acquired on a Varian INOVA 600 MHz NMR spectrometer at 27 *^∘^* C using a Carr-Purcell-Meiboom-Gill (CPMG) spin-echo pulse sequence, with a total spin-spin relaxation delay (2n*r*) of 320 ms. Water suppression was achieved by selective saturation of the water peak during the recycle delay (2s) and mixing time ™ of 150 ms. Free induction decays (FIDs) were collected into 32,000 data points with a spectral width of 8,000Hz over 64 scans. The FIDs were then zero-filled by a factor of two and multiplied by an exponential line-broadening factor of 0.5Hz prior to Fourier transformation. The diffusion-edited experiments were also carried out with bipolar pulse pair-longitudinal eddy current delay (BPP-LED) pulse sequence. The gradient amplitude was set at 35.0 G/cm with a diffusion delay of 100 ms. A total of 128 transients and 16,000 data points were collected with a spectral width of 8,000 Hz. A line-broadening factor of 1 Hz was applied to FIDs before Fourier transformation (Wang *et al*., 2004).

All NMR spectra were manually phased and baseline-corrected using VNMR 6.1C software (Varian Inc.). For CPMG spectra, each spectrum over the range of *8* 0.4–4.4 was integrated into segments of equal width (0.01 ppm). The spectrum between δ 5.2-8.5 was discarded due to the week signal of aromatic amino acids and the potential lack of association with migraine according to previous studies (Dejong *et al*.,2007; Harder *et al*.,2021). For BPP-LED data, each spectrum over the range of *8* 0.1–6.0 was segmented into integral regions of equal width (0.01ppm). The regions containing the residual signals of water (*8* 4.6–5.1) were excluded. Interval correlation shifting (icoshift) technique in MATLAB package (version 7.0) were employed to conduct peak alignment. It uses a Fast Fourier Transform (FFT) engine and a greedy algorithm that allows align all spectra simultaneously (Savorani *et al*.,2010). To accommodate these large intensity or concentration variations, the integral values of each spectrum were normalized to a constant sum of all integrals in a spectrum in order to minimize the impacts of concentration variation between samples after ppm segmentation. This total spectrum area (TSA) normalization method performed well in representing NMR spectral intensities (Emwas *et al*.,2018). Identification of metabolites in the spectra was achieved based on literatures and the Chenomx NMR Suite 4.5 (Chenomx, Calgary, Canada) and the HMDB database (http://www.hmdb.ca/). Specific compounds with multiple peaks were determined by combining the relevant ppm corresponding to the most obvious peaks in the normalized data. Figure 2B identifies and displays the major plasma metabolites. The CPMG pulse sequence was used to emphasize the resonances of small metabolites in plasma, while resonances from macromolecules were attenuated (Figure 2B). Supplementary Figure 1 shows diffusion-edited NMR spectra of plasma from each group, displaying the signals of lipid, N-acetylglycoproteins (NAc) groups of glycoproteins. Subtle differences in these spectra were observed by visual examination among groups. Following the identification of metabolites, relative concentration of metabolites could be estimated by the defined area of peak among different group. Further analysis was conducted using supervised learning statistical techniques to discriminate potential metabolic differences among the four groups.

### Statistical analysis

#### Clinical statistics

The overall statistical strategy used in this study is shown in Figure 1. The clinical variables were analyzed using R software (version 3.54). The baseline characteristics and clinical outcomes were based on the intention-to-treat (ITT) population. We omitted the cases that retained only the baseline measurement but had missing data in all clinical outcomes. If the data were normally distributed, we planned to use ANOVA to detect the differences in numerical variables and performed χ2 tests for categorical variables. If data were not normally distributed, we planned to apply the Kruskal-Wallis test. Continuous data were described as the mean (SD) with 95% CIs. Categorical data were illustrated as numbers and percentages. A one-sided test was executed for available data under hypothesis testing in superiority. A P-value <0.05 was defined as statistically significant. Clinical effect sizes were calculated by the R package “effectsize” using the cohen.d method (Ben *et al*.,2020). The standardized effect size is defined as the difference in means between two groups divided by the pooled standard deviation. The minimal important difference (MID) for the VAS was calculated and defined by the empirical work from Thorlund et al (2011), and it stands for the smallest difference in pain relief that patients regarded as important on average.

### Pattern recognition

The resulting integral data were transformed into SIMCA-P (version 14.0; Umetrics, Ume°a, Sweden) for pattern recognition analysis. Before analysis, CPMG data and BPP-LED data were Pareto-scaled, in terms of variance stabilization (Worley B, *et al*.,2013). For the purpose of discriminating differences in metabolic profiling among the groups, CPMG data and LED data were both subjected to principal component analysis (PCA). To detect and exclude the outlier of abnormal metabolic profile among included migraine patients, PCA was first performed on the normalized ^1^H NMR dataset after Pareto scaling in this study (Mickiewicz B, *et al*.,2013). A principal component (PC) score plot and loading plot were used to visualize the data. On the score plot, each point represents an individual sample, and on the loadings plot, each point indicates a single NMR spectral region. In the score plot, R^2^Y displayed the proportion of the sum of squares for the selected component, which accounts for the proportion of the variance in the responsible (y) variable explained by the regression model (Figure S2A,B) (Wang *et al*.,2004). Furthermore, OPLS-DA was performed to maximize separation and remove variance that was not related to group membership. CV-ANOVA method was also carried out for the cross-validation of OPLS-DA analysis within SIMCA-P software. Metabolites with an impact on differentiation were ranked on the basis of the variables of importance parameter (VIP) method coupled with OPLS-DA, which assesses each variable’s relative influence on the model. The metabolites with a value of VIP > 1 were selected and listed as potential biomarkers for the discrimination of model (Menni *et al*.,2017).

### Biomarker discovery and validation

To discover potential metabolite biomarkers for the effects of acupuncture, we developed a strict statistical machine learning strategy that is used for this study (see supplementary Fig. S4). Initially, the Shapiro-Wilks test was performed on raw data to check the normal distribution. Following that, OPLS-DA analysis and VIP method are conducted to achieve discrimination between groups and select potential biomarkers. After obtaining the list of potential metabolic biomarkers for migraine and acupuncture from VIP method, a Lasso regression was performed on the normalized metabolomic data using the glmnet package in R to identify the most significant metabolites determinants of change for migraine and acupuncture. The overall penalty parameter α was set to 1 to select those with the highest predicted value of metabolites in glmnet. Tenfold internal cross-validation with cv.glmnet function was applied to validate the regression model and to achieve the minimum lambda(λ), the best predict parameter for Lasso regression in the model. Through using this minimum λ yielded the most optimized model, the most relevant metabolites distinguishing migraine, acupuncture and sham acupuncture could be subsequently selected (Friedman *et al*.,2010). For the validation of potential metabolic biomarkers, ANOVA with Box-Cox transformed method was subsequently performed on these metabolites to obtain the Bonferroni-corrected P-value with the Mass package in R (Venables *et al*.,2002; Blaise *et al*.,2016). Following the analysis from VIP method, LASSO regression and ANOVA, a Venn diagram was generated to determine the overlapping metabolites selected by three aforementioned statistical methods (Supplementary Fig. S4). The overlap of statistically significant metabolites (*P* < 0.05 after Bonferroni correction) common to all three statistical methods were identified as validated biomarkers of migraine and acupuncture. Finally, receiver operating characteristic (ROC) curve analysis was performed to evaluate the diagnostic abilities of these validated biomarkers for migraine and acupuncture. GraphPad Prism version 7.0 (GraphPad Software, United States) was used to perform this ROC analysis. For the calculated area under the curve (AUC) of the biomarker, an AUC of 0.9–1.0 suggested excellent performance, and 0.8–0.89 indicated good performance, while AUC < 0.6 showed nonsignificant diagnostic performance (Haase-Fielitz *et al*.,2009). Employing this consistent statistical strategy, we were able to subsequently select and narrow the potential biomarkers from series of metabolites and pinpointed the most validated biomarkers for migraine and acupuncture (Yu Y *et al*., 2020).

### Pathway and network analysis

Ingenuity Pathway Analysis (IPA) software (IPA build version: 364062M, content version: 26127183, release date: 2015-12-12, analysis date: 2018-11-30, http://www.ingenuity.com/) (QIAGEN, Redwood City, CA, USA) was used to explore potential targeted pathways and networks related to both migraine and the effect of acupuncture in an unbiased way. The ratios of metabolites between two groups, including healthy controls vs. migraine group, acupuncture group vs. migraine group, acupuncture group vs. sham acupuncture group, were calculated and inputted into IPA software for pathway analysis of migraine and the efficacy of acupuncture. IPA’s Core Analysis module was subsequently used for pathway analysis. Fisher’s exact test was used to produce a P-value determining the probability that the link between the metabolites and the canonical pathway was explained only by chance. Canonical pathways with a *P*-value < 0.05 after Bonferroni correction were regarded as statistically significant pathways contributing to the pathology of migraine and the potential effect of acupuncture. Based on the ‘master’ network, which was developed from the Ingenuity Knowledge Base, a causal network was established to reflect observed cause-effect relationships among chemicals, protein families, complexes and biological processes. The network score was calculated using the hypergeometric distribution, with Fisher’s exact test at the right tail yielding the negative logarithm of the significant threshold. Z-score > 2 was defined as the threshold of significant activation for network analysis, disease, and function, while Z-score < 2 was defined as the threshold of significant inhibition. The score of the networks also reflects the probability of molecules gathering in this network. When the number of metabolites gathering in this network increases, the score will also increase (Krämer *et al*.,2014; Kriebel *et al*.,2016). Following the IPA analysis, the summary of significant pathways and important networks were presented in the supplementary Fig. S3.

## Data Availability

All data, models, or code used during the study are available from the corresponding author by request.

## Acknowledgements

We grateful acknowledge Prof. Xuguang Liu, Prof. Qiaofeng Wu, Prof. Zongxiang Tang, Prof. Xiaopei Shen, Prof. Fangrong Yan for their help and guide during the experiment. We thank the staff from Top Grade TCM Science Technology Laboratory of State Administration of TCM of China for the collection of blood samples of the subjects.

## Author Contributions

Jointly supervised research, F.L, S.Y, X.Y, C.G.; Conceived and designed the experiments, F.L, S.Y, X.Y.; Performed the experiments, Z.G, Q.C, L.J, X.Y; Performed statistical analysis, Z.G, C.M, C.G.; Contributed reagents, materials, or analysis tools, R.W.S, S.S, S.M, B.K, R.W, J.A, S.B.; Contributed to data exploration, Z.G, G.Y, F.G, L.J,U.M,C.G.; Wrote the paper, Z.G., X.Y, C.G. All authors discussed the results and reviewed the final manuscript.

## Supplementary information

Supplementary Fig. S1 Typical 1H NMR LED spectra of plasma samples.

Supplementary Fig. S2 Clear separation of metabolic profiles among groups.

Supplementary Fig. S3 Identification of significant pathway among three groups.

Supplementary Fig. S4 Statistical strategy for integrating different set of metabolic biomarkers from diverse statistical methods in the study.

Supplementary Table S1. Baseline characteristics.

Supplementary Table S2. Changes in plasma metabolites in LED NMR spectra between healthy controls and migraine patients

Supplementary Table S3. Changes in plasma metabolites in LED NMR spectra before and after EA treatment in migraine patients.

Supplementary Table S4. Changes in plasma metabolites in LED NMR spectra before and after sham acupuncture treatment in migraine patients

Supplementary Table S5. Changes in plasma metabolites in LED NMR spectra between EA and sham EA treatment

Supplementary Table S6. cv-anova results of OPLS-DA analysis corresponding for score plots

