## Supplemental Figure and Table 1-6 for "Metabolomics reveals reasons for the efficacy of acupuncture in migraine patients: The role of anaerobic glycolysis and mitochondrial citrate in migraine relief"

#### Supplementary data

**Figure. S1 Typical $^{1}$H NMR LED spectra of plasma samples.**

$^{1}$H NMR experiments were carried out, and Chenomx NMR Suite 4.5 software was used to identify 22 metabolites measured in a total of 50 plasma samples from 40 migraine patients and 10 healthy controls before and after EA or sham EA treatment (Table 2). A1, migraine patient; CON, healthy controls; NAc, N-acetyl methyl groups of glycoprotein； cho, choline; LDL, Low Density Lipoprotein; VLDL, Very Low Density Lipoprotein.

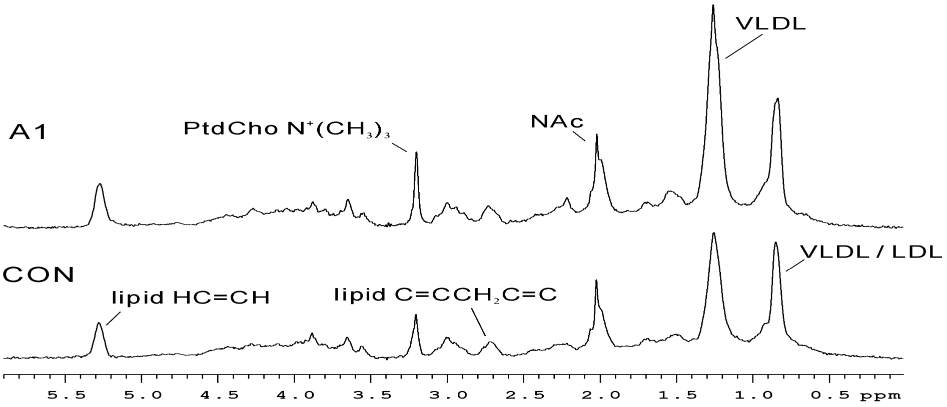

**Figure. S2 Clear separation of metabolic profiles among groups**

PCA and OPLS-DA analysis for CPMG and LED data manifested clear separation among migraine patients (red dots), healthy controls (black boxes), migraine patients after 4weeks of EA treatment (blue diamonds), and migraine patients after 4weeks of sham EA treatment (purple stars). t[1] and t[2] represent the first and second components in the PCA and OPLS-DA result, respectively. The missing samples from the EA group and Sham EA group on the score plots were excluded due to the outlier and drop out.

Separation of metabolic profiling from PCA analysis was achieved between migraine (red dots, n=40) and healthy control (black boxes, n=10) groups for CPMG data.

**(A)** Corresponding loading plots showing metabolites that may influence the separation for **(A)**

**(C)** Clear separation of metabolic profiling from OPLS-DA analysis was achieved between migraine (red dots, n=40) and healthy control (black boxes, n=10) groups for LED data.

**(D)** Corresponding loading plots showing metabolites that may influence the separation for **(A)**.

**(E)** The separation of metabolic profiling showed that EA treatment (blue diamonds, n=22) reversed the change in metabolic profiling in migraine patients (red dots, n=22) compared with healthy controls (black boxes, n=10) for LED data.

**(F)** The results showed a clear discrimination in metabolic profiling between migraine patient after EA treatment (blue diamonds, n=22) and migraine patients before EA treatment (red dots, n=22) (Table S3) for LED data.

**(G)** The results showed that migraine patient after sham EA treatment (purple stars, n=18) could not restore the change of metabolic profiling in migraine patient before sham EA treatment (red dots, n=18) compared with healthy controls (black boxes, n=10) (Table S4) for LED data.

**(H)** The result showed metabolic profiling of migraine patient before Sham EA treatment (red dots, n=18) could not be discriminated with migraine patient after Sham EA treatment (purple stars, n=18) for LED data (Table S4). 2 samples in score plot were excluded due to the outlier.

**(I)** Clear separation of metabolic profiling was discriminated between EA treatment (blue diamonds) and sham EA treatment (purple stars) for LED data (Table S5).

**A**

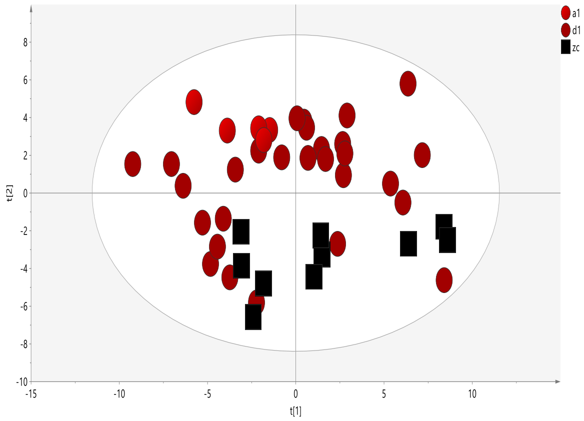

**B**

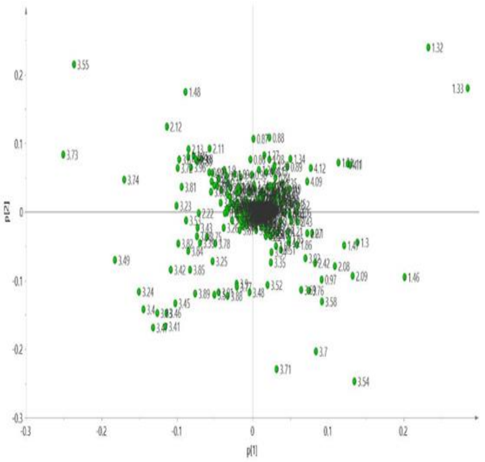

**C**

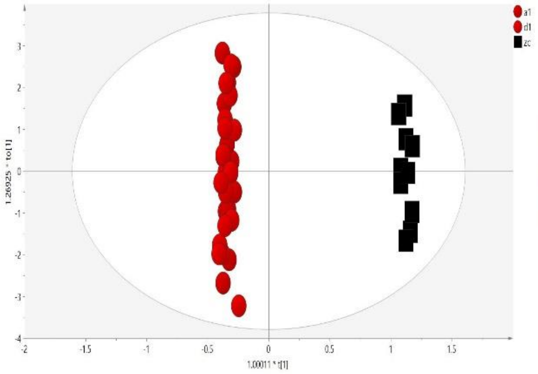

**D**

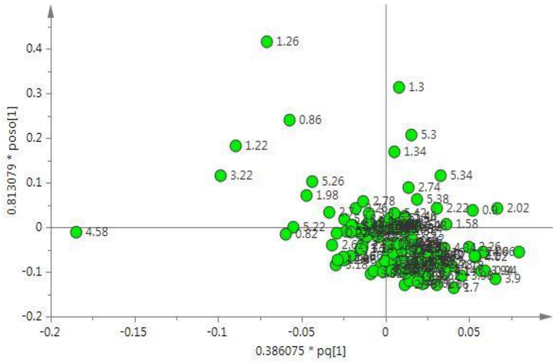

**E**

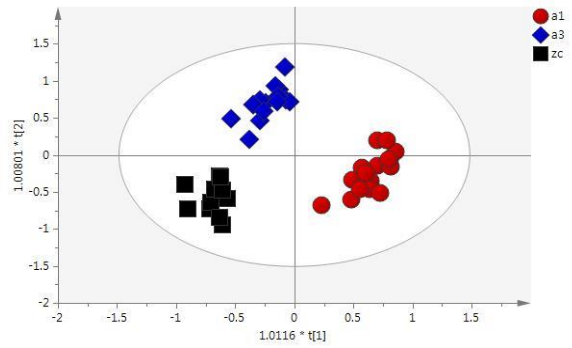

**F**

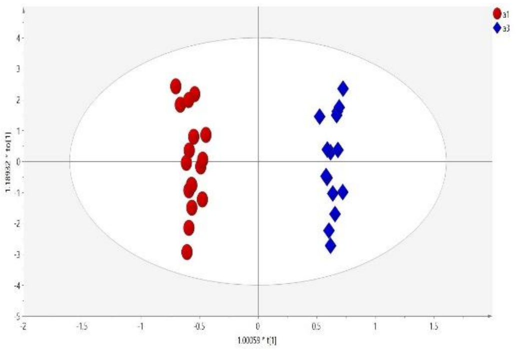

**G**

**H**

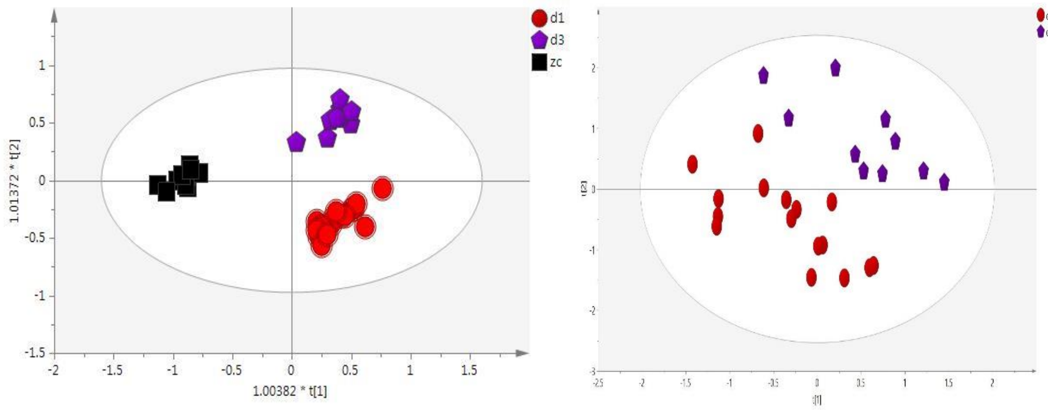

**I**

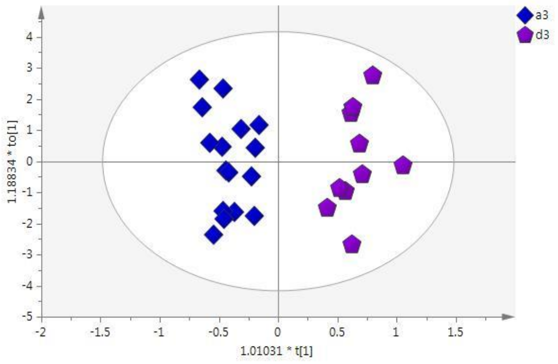

**Figure. S3 Identification of significant pathway among three groups.**

Canonical pathways between **(A)** healthy controls vs migraine patients **(B)** migraine patients before EA treatment vs migraine patients after 4weeks of EA treatment **(C)** EA vs sham EA after 4 weeks of treatment.

**(A)** Result indicates tRNA charging pathway (*P* = 0.0307) was significantly changed between healthy control and migraine patient.

**(B)** After 4 weeks EA treatment, the Pyruvate Fermentation to Lactate pathway (*P* = 0.0175) was significantly reversed in migraine patients, manifesting enhanced anaerobic glycolysis arose by EA treatment.

**(C)** Specially, tRNA Charging pathway (*P* = 0.0307) was significantly discriminated between acupuncture and sham acupuncture group.

**(D)** The network analysis result indicates the top metabolic network changed in the migraine was Carbohydrate Metabolism (score = 14).

**(E)** After 4 weeks EA treatment, the changed of carbohydrate Metabolism network which found in migraine patient was also reversed after EA treatment (score = 9).

**A**

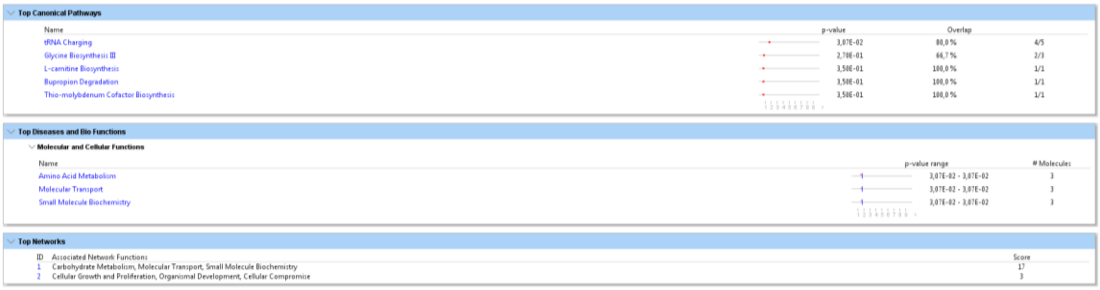

**Top Canonical Pathways**

| Name | p-value | Overlap | |
|---|---|---|---|
| tRNA Charging | 3,07E-02 | 80,0 % | 4/5 |
| Glycine Biosynthesis III | 2,70E-01 | 66,7 % | 2/3 |
| L-carnitine Biosynthesis | 3,50E-01 | 100,0 % | 1/1 |
| Bupropion Degradation | 3,50E-01 | 100,0 % | 1/1 |
| Thio-molybdenum Cofactor Biosynthesis | 3,50E-01 | 100,0 % | 1/1 |

**Top Diseases and Bio Functions**

Molecular and Cellular Functions

| Name | p-value range | # Molecules |
|---|---|---|
| Amino Acid Metabolism | 3,07E-02 - 3,07E-02 | 3 |
| Molecular Transport | 3,07E-02 - 3,07E-02 | 3 |
| Small Molecule Biochemistry | 3,07E-02 - 3,07E-02 | 3 |

**Top Networks**

| ID | Associated Network Functions | Score |
|---|---|---|
| 1 | Carbohydrate Metabolism, Molecular Transport, Small Molecule Biochemistry | 17 |
| 2 | Cellular Growth and Proliferation, Organismal Development, Cellular Compromise | 3 |

**B**

**Top Canonical Pathways**

| Name | p-value | Overlap | |
|---|---|---|---|
| Pyruvate Fermentation to Lactate | 1,75E-02 | 100,0 % | 2/2 |
| Sirtuin Signaling Pathway | 9,70E-02 | 50,0 % | 2/4 |
| HIF1α Signaling | 1,58E-01 | 100,0 % | 1/1 |
| Methylglyoxal Degradation I | 1,58E-01 | 100,0 % | 1/1 |
| Acetyl-CoA Biosynthesis III (from Citrate) | 1,58E-01 | 100,0 % | 1/1 |

**Top Upstream Regulators**

| Name | p-value | Predicted Activation |
|---|---|---|
| ICMT | 2,86E-02 | |

**Top Networks**

| ID | Associated Network Functions | Score |
|---|---|---|
| 1 | Carbohydrate Metabolism, Energy Production, Lipid Metabolism | 9 |

**C**

**Top Canonical Pathways**

| Name | p-value | Overlap | |
|---|---|---|---|
| tRNA Charging | 3,07E-02 | 80,0 % | 4/5 |
| Glycine Biosynthesis III | 2,70E-01 | 66,7 % | 2/3 |
| Leucine Degradation I | 2,70E-01 | 66,7 % | 2/3 |
| L-carnitine Biosynthesis | 3,50E-01 | 100,0 % | 1/1 |
| Bupropion Degradation | 3,50E-01 | 100,0 % | 1/1 |

**Top Diseases and Bio Functions**

**Molecular and Cellular Functions**

| Name | p-value range | # Molecules |
|---|---|---|
| Amino Acid Metabolism | 3,07E-02 - 3,07E-02 | 3 |
| Molecular Transport | 3,07E-02 - 3,07E-02 | 3 |
| Small Molecule Biochemistry | 3,07E-02 - 3,07E-02 | 3 |

**Top Networks**

| ID | Associated Network Functions | Score |
|---|---|---|
| 1 | Carbohydrate Metabolism, Molecular Transport, Small Molecule Biochemistry | 14 |
| 2 | Cellular Growth and Proliferation, Organismal Development, Cellular Compromise | 3 |

**D**

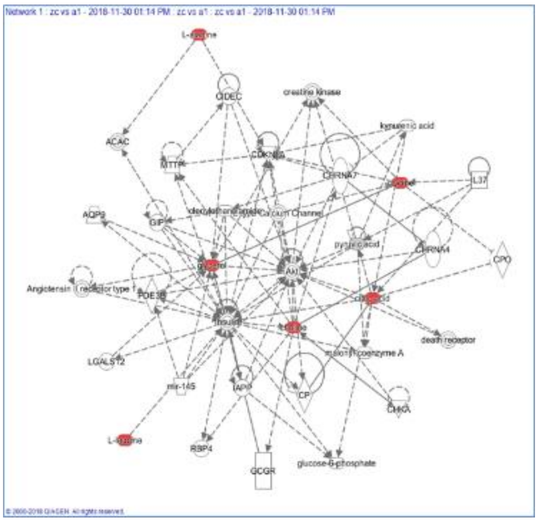

**E**

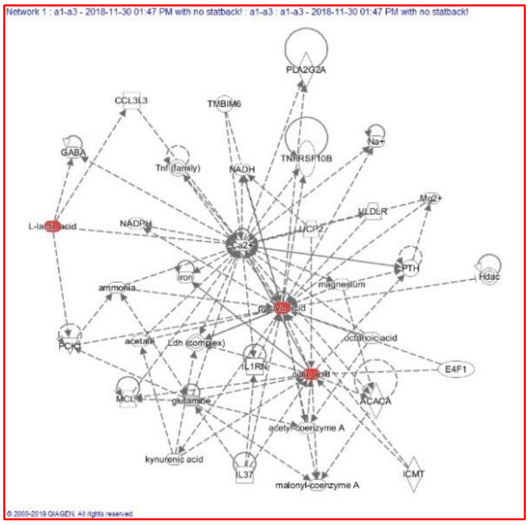

**Figure. S4 Statistical strategy for integrating different set of metabolic biomarkers from diverse statistical methods in the study.**

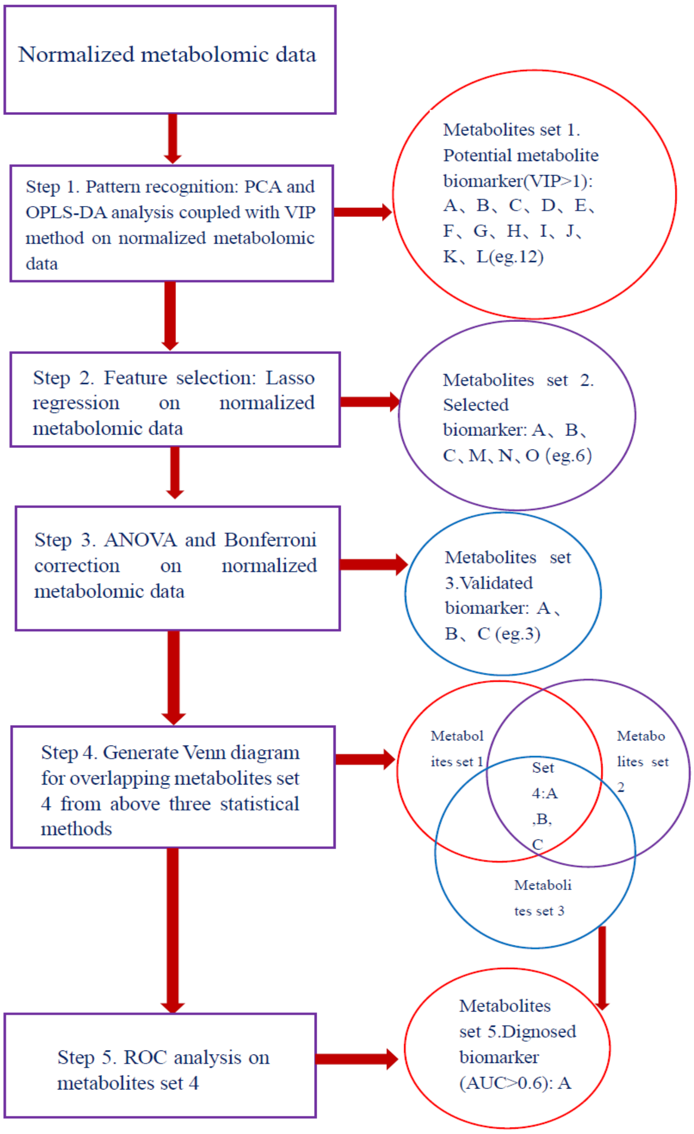

**Table S1. Baseline characteristics**

| Characteristics | EA group | Sham EA group | Statistical value | P value |
|---|---|---|---|---|
| No. of patients (n) | 22# | 18# | | |
| No. of women, n ( % ) | 22(100%) | 18(100%) | | |
| Age (y), mean (SD) | 30.14(9.14) | 27.87(7.34) | 0.74 | 0.46 |
| Height (cm), mean (SD) | 158(4.83) | 159.25(5.48) | -0.66 | 0.51 |
| Weight (kg), mean (SD) | 52.85(5.93) | 50.06(5.38) | 1.34 | 0.19 |
| Course of disease (mo), | 118.07(103.78) | 95.8(42.97) | 0.74 | 0.46 |

mean

### The baseline characteristics were based on the intention-to-treat (ITT) population. We omitted the cases that retained only the baseline measurement but had missing data for all clinical outcomes.mo, month.

**Table S2. Changes in plasma metabolites in LED NMR spectra between healthy controls and migraine patients**

| **Metabolites** | **Peak Regions** | **Con (n=10) mean(sd)** | **Mig (n=40) mean(sd)** | **Direction of effect Mig vs Con** | **adjP Mig vs Con** |
|---|---|---|---|---|---|
| NAC | 2.06 | 162.50(8.49) | 153.21(6.66) | ↓↓ | 0.00082** |
| Lipid | 1.54 | 127.64(4.28) | 126.59(4.29) | ↓ | 0.48 |
| VLDL | 1.26 | 661.75(56.94) | 684.60(63.80) | ↑ | 0.31 |
| UFA (Unsaturated fatty acids) | 5.3 | 134.99(13.73) | 132.47(16.40) | ↓ | 0.66 |
| LDL/VLDL | 0.86 | 481.36(30.70) | 494.43(33.69) | ↑ | 0.26 |
| Ptdcho | 3.22 | 202.43(26.06) | 225.52(31.68) | ↑↑ | 0.035* |

The up or down arrow ( ↑ / ↓ ) indicates whether the structure showed a signal increase or decrease, respectively. **P<0.05; **P<0.01*. Abbreviations: Con, healthy control; Mig, migraine patients from the EA and Sham EA groups before treatment; adjp, p value after Bonferroni correction.

**Table S3. Changes in plasma metabolites in LED NMR spectra before and after EA treatment in migraine patients**

| **Metabolites** | **Con (n=10) mean(sd)** | **Mig (n=22) mean(sd)** | **Direction of effect Con vs Mig** | **adjP Mig vs Con** | **EA mean(sd)** | **Direction of effect Con vs EA** | **adjP EA vs Con** | **Direction of effect EA vs Mig** | **adjP EA vs Mig** |
|---|---|---|---|---|---|---|---|---|---|
| NAC | 162.50(8.49) | 152.63(8.26) | ↓↓ | 0.0031** | 150.95(4.59) | ↓↓ | 0.00073** | ↓ | 0.79 |
| Lipid | 127.64(4.28) | 125.58(4.06) | ↓ | 0.38 | 125.97(3.55) | ↓ | 0.54 | - | 0.96 |
| VLDL | 661.75(56.94) | 684.74(66.56) | ↑ | 0.61 | 659.13(61.18) | ↓ | 0.99 | ↓ | 0.48 |
| UFA(Unsaturated fatty acids) | 134.99(13.73) | 133.37(15.59) | ↓ | 0.96 | 128.26(17.50) | ↓ | 0.54 | ↓ | 0.64 |
| LDL/VLDL | 481.36(30.70) | 494.04(32.84) | ↑ | 0.60 | 480.28(36.45) | - | 0.99 | ↓ | 0.49 |
| Ptdcho | 202.43(2 | 228.81(3 | ↑ | 0.073 | 221.09(3 | ↑ | 0.28 | ↑ | 0.7 |

| | | | | | | | | | |
|---|---|---|---|---|---|---|---|---|---|
| | 6.05) | 2.06) | | | 0.48) | | | | 5 |

The up or down arrow ( ↑ / ↓ ) indicates whether the structure showed a signal increase or decrease, respectively. **P<0.05; **P<0.01*. Mig, migraine patients in EA group before treatment; EA, migraine patients in the EA group after 4 weeks of EA treatment. Con, healthy control. adjp, p value after Bonferroni correction.

**Table S4. Changes in plasma metabolites in LED NMR spectra before and after sham acupuncture treatment in migraine patients**

| Metabolites | Con mean(sd) (n = 10) | Mig mean(sd) (n = 18) | Direction of effect Con vs Mig | adjP Mig vs Con | Sham EA mean(sd) | Direction of effect Sham EA vs Con | adjP Sham EA vs Con | Direction of effect Sham EA vs Mig | adjP Sham EA vs Mig |
|---|---|---|---|---|---|---|---|---|---|
| NAC | 162.50(8.50) | 153.84(4.60) | ↓↓ | 0.0048** | 153.68(6.70) | ↓↓ | 0.0104* | - | 0.99 |
| Lipid | 127.64(4.28) | 127.66(4.39) | - | 0.99 | 128.74(4.60) | ↑ | 0.84 | ↑ | 0.82 |
| VLDL | 661.75(56.94) | 684.44(62.90) | ↑ | 0.61 | 704.46(62.43) | ↑ | 0.26 | ↑ | 0.70 |
| UFA(Unsaturated fatty acids) | 134.99(13.73) | 131.51(17.68) | ↓ | 0.85 | 136.13(16.30) | ↑ | 0.98 | ↑ | 0.76 |
| LDL/VLDL | 481.36(30.69) | 494.86(35.64) | ↑ | 0.63 | 515.24(46.35) | ↑ | 0.11 | ↑ | 0.37 |
| Ptdcho | 202.43(26.05) | 222.02(31.94) | ↑ | 0.24 | 226.93(33.60) | ↑ | 0.17 | ↑ | 0.92 |

The up or down arrow ( ↑ / ↓ ) indicates whether the structure showed a signal increase or decrease, respectively. **P<0.05; **P<0.01*. Mig, migraine patients in sham EA group before treatment; Sham EA, migraine patients in the sham EA group after 4 weeks of sham EA treatment. Con, healthy control. adjp, p value after Bonferroni correction.

**Table S5. Changes in plasma metabolites in LED NMR spectra between EA and sham EA treatment**

| Metabolites | EA mean(sd) | Sham EA mean(sd) | Direction of effect EA vs Sham EA | adjP EA vs Sham EA | Coefficient EA vs Sham EA |
|---|---|---|---|---|---|
| NAC | 151.24(4.58) | 153.69(6.70) | ↓ | 0.26 | 0.80 |
| Lipid | 125.98(3.43) | 128.744(4.60) | ↓ | 0.10 | 0.42 |
| VLDL | 658.16(59.24) | 694.67(57.16) | ↓ | 0.13 | n.a. |
| UFA(unesteri | 127.92(16.97) | 133.73(15.53) | ↓ | 0.39 | n.a. |

| | | | | | |
|---|---|---|---|---|---|
| fied fatty acids) | | | | | |
| LDL/VLDL | 478.09(36.29) | 507.32(46.20) | ↓ | 0.094 | 1.11 |
| Ptdcho | 219.54(30.09) | 224.16(33.52) | ↓ | 0.74 | n.a. |

The up or down arrow (↑/↓) indicates whether the metabolite showed a signal increase or decrease, respectively. **P<0.05; **P<0.01*. The coefficient was calculated by Lasso regression. EA, migraine patients in the EA group after 4 weeks of EA treatment; Sham EA, migraine patients in the Sham EA group after 4 weeks of Sham EA treatment.

**Table S6. Cross-validation results for OPLS-DA analysis using CV-ANOVA**

| Group | F value | P value |
|---|---|---|
| **CPMG data** | | |
| Mig vs con | 8.56 | 5.01158e-005 |
| Mig vs EA | 2.55 | 0.031 |
| Mig vs sham EA | 0 | 1 |
| EA vs sham EA | 4.18 | 0.0145 |
| **LED data** | | |
| Mig vs con | 6.43 | 1.67389e-006 |
| Mig vs EA | 15.41 | 1.92117e-007 |
| Mig vs sham EA | 3.417 | 0.0266 |
| EA vs sham EA | 8.09923 | 0.000195154 |

**P<0.05; **P<0.01*. The F and P values were calculated by CV-ANOVA in SIMICA-P software. Mig, migraine patients before treatment; EA, migraine patients in EA group after 4 weeks of acupuncture treatment; Sham EA, migraine patients in the sham EA group after 4 weeks of sham EA treatment. Con, healthy control.
